# Lifetime non-relational traumatic experiences and biological ageing

**DOI:** 10.1101/2025.08.15.25333766

**Authors:** Thole H Hoppen, Nexhmedin Morina, Monica Aas, Julian Mutz

## Abstract

**Background:** Exposure to non-relational trauma, such as serious accidents, war or life-threatening illness, is linked to poor mental and physical health. Its relationship with multiple biological ageing domains, however, remains underexplored.

**Objective:** To examine associations between non-relational trauma and markers of biological ageing, and to assess whether associations vary by trauma burden, trauma type and sex.

**Methods:** We analysed UK Biobank data from 152,863 participants (mean age = 56.40 years; 56.50% female). Lifetime exposure to six non-relational traumatic experiences was assessed. Biological ageing markers included metabolomic age (MileAge) delta, a metabolomic mortality profile score, frailty index, leukocyte telomere length and grip strength. Regression models, adjusted for demographic and socioeconomic covariates, estimated associations between trauma and biological ageing markers. We also examined dose-response, trauma type-specific and sex-specific associations.

**Results:** Non-relational trauma was associated with a metabolite-predicted age exceeding chronological age, elevated metabolomic mortality scores and greater frailty. All non-relational trauma types were associated with greater frailty, with a clear dose-response pattern and with the strongest association observed for life-threatening illness. There was no evidence of associations with telomere length, and mixed findings for grip strength. Several associations differed by sex, for example overall trauma burden was more strongly associated with greater frailty in females compared to males.

**Conclusions:** Lifetime non-relational trauma was associated with older biological ageing profiles, with the strongest associations with frailty. These findings support the notion that non-relational trauma exposure is associated with long-term health status, underscoring the need for mitigating ageing-related health decline in trauma-exposed populations.

## Introduction

Non-relational traumatic experiences, such as serious accidents, war or life-threatening illness, can have profound and lasting effects on both mental and physical health (Bourassa & Sbarra, 2024; Hoppen, Priebe, Vetter, & Morina, 2021; Kessler et al., 2017). Unlike relational trauma, where the perpetrator is known to the trauma survivor, non-relational trauma either involves an unknown perpetrator (e.g., combat) or no perpetrator (e.g., life-threatening illness). Relational and non-relational trauma have both been linked to increased risks of mental disorders, chronic diseases and morbidity (Bourassa & Sbarra, 2024; Hoppen et al., 2021; Kessler et al., 2017). Both have also been linked to an increased risk of premature mortality (Grummitt et al., 2021) and to metabolic changes (Mellon, Gautam, Hammamieh, Jett, & Wolkowitz, 2018).

One proposed mechanism linking trauma and adverse health outcomes is accelerated biological ageing, which refers to the progressive accumulation of molecular and cellular damage and the decline of physiological integrity. Biological ageing can be assessed using various molecular, clinical and functional markers (Furrer & Handschin, 2025; Pickhardt et al., 2025; Teschendorff & Horvath, 2025), including ageing clocks, frailty indices, telomere length and grip strength. Each of these measures provides complementary insights into processes of systemic decline. Metabolomics, in particular, is a powerful tool to capture metabolic perturbations shaped by both genetic and environmental exposures (Fiehn, 2002; Huang et al., 2025). Frailty indices reflect the accumulation of age-related health deficits and are associated with psychosocial stressors (Lee et al., 2023). Similarly, telomere attrition and muscular decline have been linked to chronic stress and traumatic experiences (Duchowny et al., 2024; Epel et al., 2004). However, associations between non-relational trauma and multiple dimensions of biological ageing remain comparatively underexplored.

Sex differences in life expectancy, health outcomes, and stress reactivity are well-documented. On average, females live longer than males despite reporting higher rates of chronic illness and disability in later life (Austad & Fischer, 2016). Females also experience higher rates of certain types of traumas, such as sexual assault, whereas males are more often exposed to physical assault and combat-related trauma (Tolin & Foa, 2006). Furthermore, biological ageing processes may differ by sex (Hägg & Jylhävä, 2021; Mutz & Lewis, 2021). These observations highlight the importance of examining biological sex as a potential moderator in the relationship between trauma exposure and biological ageing. Yet, few studies have tested whether associations of non-relational trauma with biological ageing differ between males and females.

Although there is growing interest in the biological embedding of trauma, much of the existing research has focused on relational trauma or adversity more broadly or has relied on small samples (Nelles-McGee, Khoury, Kenny, Joshi, & Gonzalez, 2022). Studies also often examined single ageing markers in isolation, limiting insights into the broader cumulative impact of severe trauma. Building on our prior study that identified associations between childhood and adulthood adversity and diverse markers of ageing (Aas et al., 2025), we sought to investigate associations between non-relational trauma and several biological ageing markers in a large, population-based cohort.

Using data from the UK Biobank, we examined whether lifetime exposure to non-relational traumatic experiences is associated with metabolomic ageing, frailty, telomere length and grip strength. We also tested whether associations varied by the number of distinct trauma exposures (“dose-response”), by type of traumatic experience any by sex. We hypothesised that non-relational trauma would be associated with older biological ageing profiles in both sexes, with stronger associations for greater trauma burden.

## Methods

### Study population

The UK Biobank recruited over 500,000 individuals from England, Scotland and Wales. At baseline (2006-2010), participants completed health and sociodemographic questionnaires, underwent physical examinations and provided biological samples. Exposure to adversity and trauma was assessed via the online follow-up Mental Health Questionnaire (MHQ) administered between 2016 and 2017 (Davis et al., 2020).

### Non-relational trauma

Lifetime exposure to non-relational trauma was assessed using a six-item checklist (Frissa et al., 2016). Items included: “In your life, have you … 1) been in a serious accident that you believed to be life-threatening at the time; 2) been involved in combat or exposed to a war-zone (either in the military or as a civilian); 3) been diagnosed with a life-threatening illness; 4) been attacked, mugged, robbed, or been the victim of a physically violent crime; 5) witnessed a sudden violent death (e.g., murder, suicide, aftermath of an accident); 6) been a victim of a sexual assault, whether by a stranger or someone you knew.” Response options were “Never”, “Yes, but not in the last 12 months” and “Yes, within the last 12 months.” Participants were classified as having experienced non-relational trauma (yes/no) if they responded “Yes, but not in the last 12 months” to at least one item. Those who responded “Yes, within the last 12 months” were excluded from analyses as these events occurred after the biomarker data were collected. A sum score (range: 0-6) was also derived to reflect overall trauma burden.

### Metabolomic age (MileAge) delta

Nuclear magnetic resonance (NMR) spectroscopy metabolomic biomarkers were quantified in non-fasting plasma samples using the Nightingale Health platform, which measures 249 biomarkers (168 absolute concentrations and 81 derived ratios) via a high-throughput protocol (Würtz et al., 2017). Technical variation was removed using the ‘ukbnmr’ R package (algorithm v2) (Ritchie et al., 2023). In a prior study (Mutz, Iniesta, & Lewis, 2024), we developed a metabolomic ageing clock using a Cubist rule-based regression model.

Individual age predictions were aggregated across the ten test sets of the outer nested cross-validation loop to avoid overfitting. Metabolomic age (MileAge) delta was defined as the difference between metabolite-predicted and chronological age, with positive values indicating an older biological age profile.

### Metabolomic mortality profile score

In a separate study, we developed a metabolomic mortality profile score (Zhang et al., 2024). Complementing the 249 biomarkers directly provided by Nightingale Health, 76 additional lipid, cholesterol and fatty acid ratios were derived (Ritchie et al., 2023), yielding a total of 325 biomarkers. A Least Absolute Shrinkage and Selection Operator (LASSO) Cox proportional hazards model predicting time to mortality was developed in English and Welsh participants (*N* = 234,553). The metabolomic mortality profile score was calculated for Scottish participants (*N* = 15,788) as a linear combination of the 54 biomarkers with non-zero coefficients in the LASSO Cox model weighted by their estimated log hazard ratios. Higher scores indicate an elevated mortality risk.

### Frailty index

A frailty index was derived from 49 health deficits reported via touch-screen questionnaires or during nurse-led interviews, in accordance with the following criteria: indicators of poor health, more common in older individuals, neither rare nor universal, covering multiple areas of functioning and available for ≥ 80% of participants (Williams, Jylhävä, Pedersen, & Hägg, 2019). Variables were dichotomised (deficit absent = zero; deficit present = one) or mapped to a score between zero and one, if ordinal. The sum of deficits was divided by the total number of possible deficits, resulting in frailty index values between zero and one, with higher values indicating greater frailty (Mutz, Choudhury, Zhao, & Dregan, 2022). Participants missing ≥ 10 of 49 variables were excluded (Williams et al., 2019).

### Telomere length

Leukocyte telomere length was measured using a quantitative polymerase chain reaction (qPCR) assay that expresses telomere length as the ratio of the telomere repeat copy number (T) relative to a single-copy gene (S) encoding haemoglobin subunit beta (Codd et al., 2022). T/S ratio is proportional to average telomere length (Lai, Wright, & Shay, 2018). Measurements were adjusted for operational and technical parameters (PCR machine, staff member, enzyme batch, primer batch, temperature, humidity, primer batch × PCR machine, primer batch × staff member, A260/A280 ratio of the DNA sample and A260/A280 ratio squared), log*_e_* transformed and *Z*-standardised.

### Grip strength

Maximal grip strength (kilogram force) was measured using a Jamar J00105 hydraulic hand dynamometer (range: 0-90 kilogram). We used the maximal value from the participant’s self-reported dominant hand. If handedness data were unavailable, the highest value was used.

### Covariates

Potential confounders included chronological age, sex, highest educational or professional qualification, gross annual household income, ethnicity and neighbourhood deprivation (Townsend deprivation index).

### Exclusion criteria

Participants with missing or “prefer not to answer” response to any non-relational trauma item were excluded from the binary and sum score trauma variables. For item-specific analyses, only those missing data for the relevant item were excluded. For categorical covariates, “do not know’, “prefer not to answer” and missing values were coded as a separate missing category. For analyses of MileAge delta, we applied exclusions as per the original study (Mutz et al., 2024): women pregnant or uncertain of pregnancy status (due to altered metabolite profiles); participants with discordant genetic and self-reported sex (indicating possible data quality issues); and participants with missing or extreme metabolite values (>4× the interquartile range from the median).

### Statistical analyses

All analyses were performed in R (version 4.3.0). Descriptive statistics included means and standard deviations or counts and percentages.

Associations between non-relational trauma (exposure) and each outcome (MileAge delta, metabolomic mortality profile score, frailty index, telomere length and grip strength) were estimated using ordinary least squares regression. All outcomes were scaled (mean = 0, standard deviation = 1) to allow for direct comparisons of association estimates. Telomere length and grip strength were reverse coded prior to analysis. We examined binary (yes/no), unweighted sum score and ordinal (based on the sum score, with levels 5-6 collapsed due to sparse data) exposure definitions. We further examined associations with exposure to multiple (at least two) traumatic events. For each outcome, we fitted a model adjusted for chronological age and sex (Model 1) and model also adjusted for education, income, ethnicity and neighbourhood deprivation (Model 2). *P*-values were corrected for multiple testing using the Benjamini-Hochberg method, with a two-tailed test and a false discovery rate of 5%. To investigate whether specific types of traumas were associated with biological ageing, we further examined associations for each questionnaire item, using the same modelling strategy. Finally, to explore potential sex differences, we estimated sex-stratified associations and assessed potential sex-by-trauma interactions by adding cross-product terms for both the trauma sum score and each individual questionnaire item.

### Sensitivity analysis

We performed a sensitivity analysis excluding the item ‘… been a victim of a sexual assault, whether by a strange or someone you knew’ from the non-relational trauma sum score to avoid potential overlap with our prior study (Aas et al., 2025). We then re-ran all analyses using the modified sum score (range: 0-5), including overall associations, sex-stratified models and models testing the interaction between the trauma sum score and sex.

## Results

Amongst 152,863 participants (mean age = 56.40 years, SD = 7.73; 56.50% female, 77,057 reported lifetime exposure to non-relational trauma (Table S1). Analytical sample sizes for each biological ageing marker are reported in Table S2. Non-relational trauma (yes/no) was associated with a metabolite-predicted age exceeding chronological age (*β* = 0.047, 95% CI 0.032-0.062, *p* < 0.001), higher metabolomic mortality profile scores (*β* = 0.102, 95% CI 0.051-0.153, *p* < 0.001) and higher frailty index values (*β* = 0.298, 95% CI 0.290-0.307, *p* < 0.001) (Figure 1A). There was no evidence of associations with telomere length or grip strength (Table 1).

**Figure 1.**
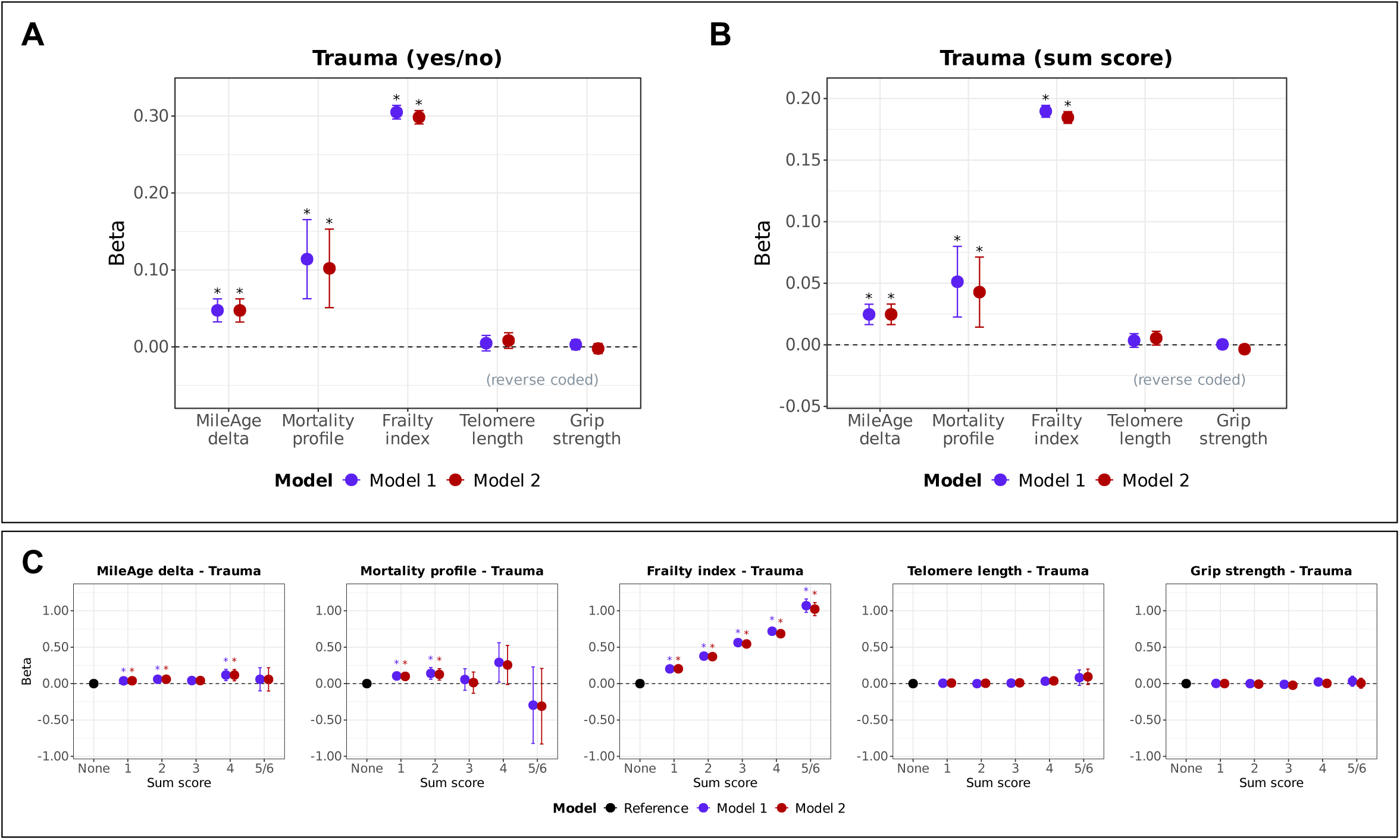
**(A) Trauma (yes/no) and ageing markers.** Associations between lifetime exposure to non-relational trauma and ageing markers (MileAge delta, frailty index, telomere length [reverse coded] and grip strength [reverse coded]). Asterisks indicate statistically significant associations, after correcting p-values for multiple testing using the Benjamini–Hochberg procedure (across ageing markers and models). **(B) Trauma (sum scores) and ageing markers.** Associations between lifetime exposure to non-relational trauma and ageing markers (MileAge delta, frailty index, telomere length [reverse coded] and grip strength [reverse coded]). Asterisks indicate statistically significant associations, after correcting p-values for multiple testing using the Benjamini–Hochberg procedure (across ageing markers and models). **(C) Trauma (sum scores, categorical) and ageing markers.** Associations between lifetime exposure to non-relational trauma and ageing markers (MileAge delta, frailty index, telomere length [reverse coded] and grip strength [reverse coded]). Asterisks indicate statistically significant associations, after correcting p-values for multiple testing using the Benjamini–Hochberg procedure (across exposure levels and models, separately for each ageing marker). **(A-C)** Estimates shown are ordinary least squares regression beta coefficients and 95% confidence intervals. Model 1–adjusted for chronological age and sex; Model 2–adjusted for chronological age, sex, ethnicity, education, income and neighbourhood deprivation. Sample sizes reported in Table S2.

**Table 1.** Associations between non-relational trauma and ageing markers (binary)

|  | Model 1 (adj. age and sex) |  |  |  | Model 2 (full adjustment) |  |  |  |
| --- | --- | --- | --- | --- | --- | --- | --- | --- |
| | $\beta$ | 95% CI | | $p$ | $\beta$ | 95% CI | | $p$ |
| Ageing marker |  |  |  |  |  |  |  |  |
| MileAge delta | 0.048 | 0.033 | 0.062 | <0.001 | 0.047 | 0.032 | 0.062 | <0.001 |
| Metabolomic profile | 0.114 | 0.063 | 0.166 | <0.001 | 0.102 | 0.051 | 0.153 | <0.001 |
| Frailty index | 0.305 | 0.296 | 0.314 | <0.001 | 0.298 | 0.290 | 0.307 | <0.001 |
| Telomere length | 0.005 | -0.005 | 0.015 | 0.411 | 0.008 | -0.002 | 0.018 | 0.153 |
| Grip strength | 0.003 | -0.004 | 0.010 | 0.411 | -0.002 | -0.009 | 0.004 | 0.533 |

*Note:* CI = confidence interval. Model 1—adjusted for chronological age and sex; Model 2—adjusted for chronological age, sex, ethnicity, highest educational/professional qualification, annual gross household income and Townsend deprivation index. *P*-values shown are corrected for multiple testing using the Benjamini–Hochberg procedure (across ageing markers and models). Sample sizes reported in Table S2.

### Dose-response associations with trauma burden

Exposure to multiple (at least two) non-relational traumas was associated with a MileAge exceeding chronological age, higher metabolomic mortality profile scores and higher frailty index values (Figure S1). Exposure to multiple non-relational traumas was nominally associated with higher grip strength after full adjustment (Table S3). Analyses of the sum scores mirrored the results of the binary exposures (Figure 1B). There was no evidence of an association with telomere length or grip strength after multiple testing correction (both *p* = 0.073) (Table 2). Sum scores of one, two and four were associated with a metabolite-predicted age exceeding chronological age (Table S4). Sum scores of one and two were also associated with higher metabolomic mortality profile scores. There was a strong dose-response association between the number of non-relational trauma exposures and higher frailty index values (ranging from *β* = 0.204, 95% CI 0.194-0.214, *p* < 0.001 to *β* = 1.024, 95% CI 0.933-1.114, *p* < 0.001) (Figure 2C). There was no evidence that any number of non-relational trauma exposures was associated with telomere length. Finally, sum scores of three were nominally associated with higher grip strength (*β* = -0.022, 95% CI -0.040 to -0.005, *p* = 0.114).

**Figure 2.**
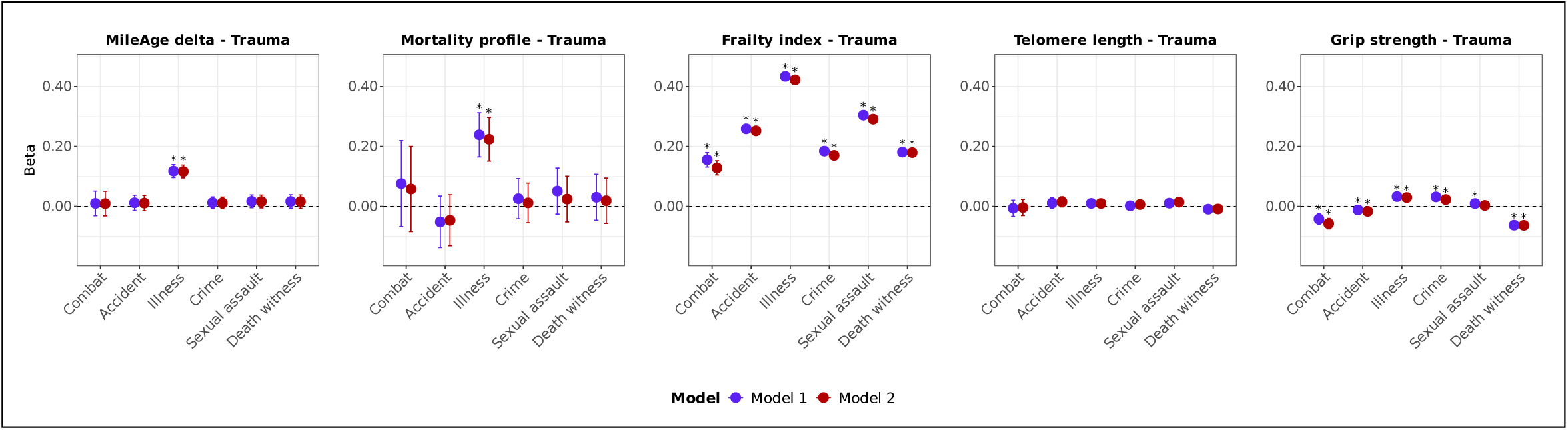
Trauma (item-specific) and ageing markers. Associations between specific trauma items (lifetime exposure to non-relational trauma) and ageing markers (MileAge delta, frailty index, telomere length [reverse coded] and grip strength [reverse coded]). Estimates shown are ordinary least squares regression beta coefficients and 95% confidence intervals. Model 1–adjusted for chronological age and sex; Model 2–adjusted for chronological age, sex, ethnicity, education, income and neighbourhood deprivation. Asterisks indicate statistically significant associations, after correcting p-values for multiple testing using the Benjamini–Hochberg procedure (across trauma items and models, separately for each ageing marker). Sample sizes reported in Table S2.

**Table 2.** Associations between non-relational trauma and ageing markers (sum score) Model 1 (adj. age and sex) Model 2 (full adjustment)

|  | Model 1 (adj. age and sex) |  |  |  | Model 2 (full adjustment) |  |  |  |
| --- | --- | --- | --- | --- | --- | --- | --- | --- |
| | $\beta$ | 95% CI | | $p$ | $\beta$ | 95% CI | | $p$ |
| Ageing marker |  |  |  |  |  |  |  |  |
| MileAge delta | 0.025 | 0.016 | 0.033 | <0.001 | 0.025 | 0.016 | 0.033 | <0.001 |
| Metabolomic profile | 0.051 | 0.023 | 0.080 | <0.001 | 0.043 | 0.014 | 0.071 | 0.005 |
| Frailty index | 0.190 | 0.185 | 0.194 | <0.001 | 0.185 | 0.180 | 0.189 | <0.001 |
| Telomere length | 0.004 | -0.002 | 0.009 | 0.238 | 0.005 | 0.000 | 0.011 | 0.073 |
| Grip strength | 0.000 | -0.003 | 0.004 | 0.842 | -0.003 | -0.007 | 0.000 | 0.073 |

*Note:* CI = confidence interval. Model 1—adjusted for chronological age and sex; Model 2—adjusted for chronological age, sex, ethnicity, highest educational/professional qualification, annual gross household income and Townsend deprivation index. *P*-values shown are corrected for multiple testing using the Benjamini–Hochberg procedure (across ageing markers and models). Sample sizes reported in Table S2.

### Specific types of non-relational trauma

Diagnosis of a life-threatening illness was the only non-relational trauma associated with a MileAge exceeding chronological age (*β* = 0.117, 95% CI 0.095-0.138, *p* < 0.001) and higher mortality profile scores (*β* = 0.224, 95% CI 0.151-0.297, *p* < 0.001) (Table S5). All non-relational trauma exposures were associated with higher frailty index values, with the strongest association observed for life-threatening illness (*β* = 0.422, 95% CI 0.410-0.435, *p* < 0.001) (Figure 2). No associations between non-relational trauma and telomere length were statistically significant after multiple testing correction (*p* ≥ 0.371). We identified mixed associations between non-relational trauma and grip strength. Life-threatening illness and having been a victim of crime were both associated with weaker grip strength. However, combat or warzone exposure, serious accidents or witnessing a sudden violent death were all associated with higher grip strength.

### Sex differences

Amongst 152,863 participants, 46.29% of females (39,950 out of 86,311) and 55.76% of males (37,107 out of 66,552) reported lifetime exposure to non-relational trauma (Table S6). Males had higher trauma sum scores than females (Table S7). Combat exposure was more common amongst males (6.1%) than females (1.5%), whereas sexual assault was more prevalent amongst females (20.6%) than males (7.6%). Analytical sample sizes are shown in Table S7.

In sex-stratified analyses, associations between non-relational trauma and metabolomic ageing were comparable in males and females (Figure 3; Table S8). Associations with frailty were stronger in females (*β* = 0.203, 95% CI 0.196-0.210, *p* < 0.001) than in males (*β* = 0.167, 95% CI 0.160-0.173, *p* < 0.001), with a statistically significant interaction (*β* _interaction_ = -0.035, 95% CI -0.044 to -0.025, *p* < 0.001) (Table S9). After full adjustment, non-relational trauma (sum score) was associated with lower grip strength in females (*β* = 0.005, 95% CI 0.001-0.009, *p* = 0.037) and with higher grip strength in males (*β* = -0.009, 95% CI -0.015 to -0.003, *p* = 0.005). However, the interaction was not statistically significant (*β* _interaction_ = - 0.007, 95% CI -0.014 to 0.000, *p* = 0.146).

**Figure 3.**
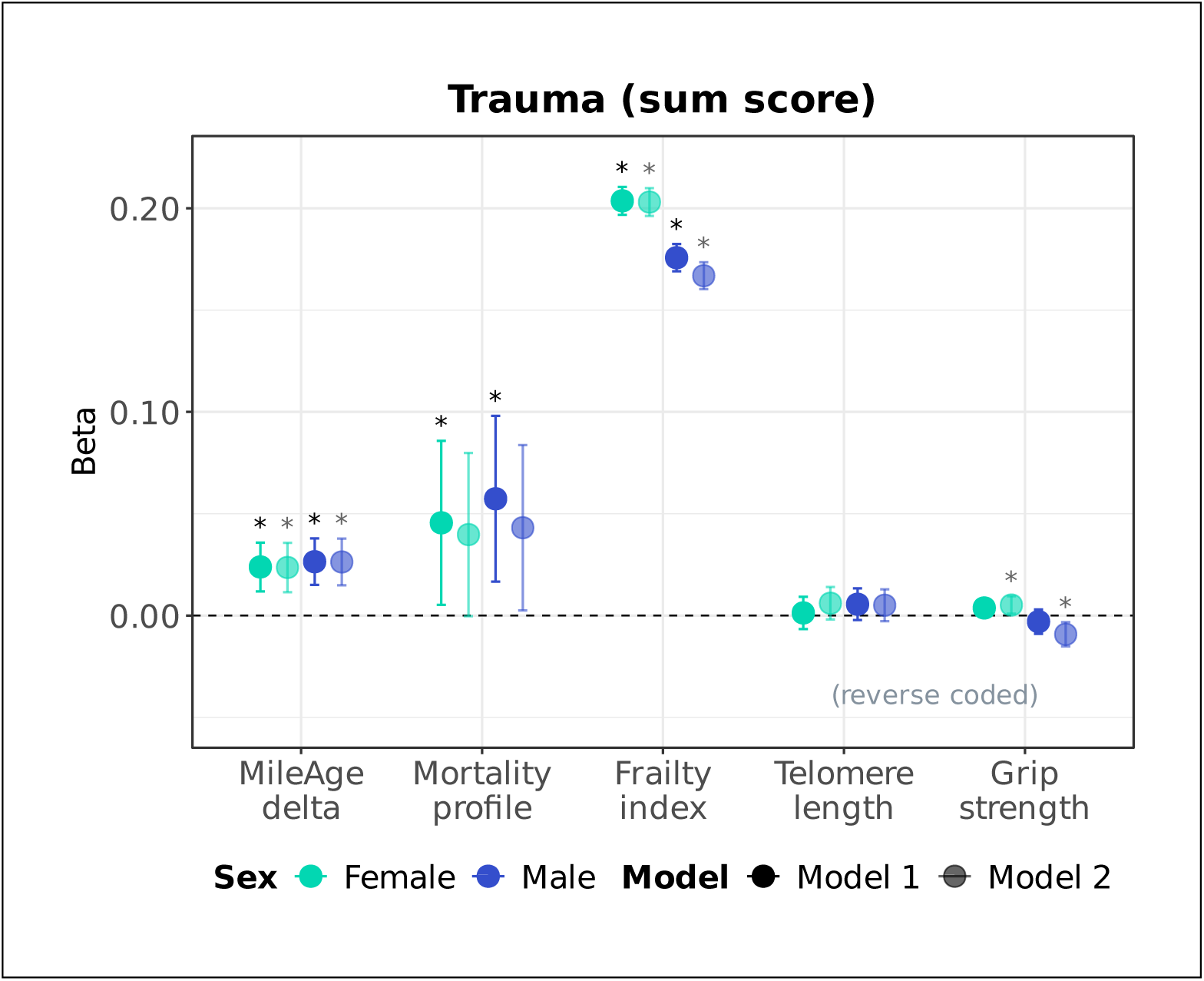
Sex-stratified associations between trauma (sum scores) and ageing markers. Sex-stratified associations between lifetime exposure to non-relational trauma and ageing markers (MileAge delta, frailty index, telomere length [reverse coded] and grip strength [reverse coded]). Estimates shown are ordinary least squares regression beta coefficients and 95% confidence intervals. Model 1–adjusted for chronological age; Model 2–adjusted for chronological age, ethnicity, highest educational/professional qualification, annual gross household income and neighbourhood deprivation. Asterisks indicate statistically significant associations, after correcting *p*-values for multiple testing using the Benjamini– Hochberg procedure (across ageing markers and models, separately for each sex). Sample sizes reported in Table S7.

Life-threatening illness was more strongly associated with MileAge delta in males (*β* = 0.150, 95% CI 0.119-0.181, *p* < 0.001) than in females (*β* = 0.096, 95% CI 0.066-0.126, *p* < 0.001) (Figure 4; Table S10), though the interaction was not statistically significant (*p* _interaction_ > 0.999) (Table S11). Sexual assault was associated with a metabolite-predicted age exceeding chronological age only in females (*β* = 0.034, 95% CI 0.008-0.059, *p* = 0.022). The metabolomic mortality profile score was also more strongly associated with life-threatening illness in males (*β* = 0.275, 95% CI 0.165-0.384, *p* < 0.001) than in females (*β* = 0.183, 95% CI 0.085-0.281, *p* < 0.001), although the interaction test was again not statistically significant (*p* _interaction_ = 0.305). Life-threatening illness was more strongly associated with frailty in males (*β* = 0.460, 95% CI 0.442-0.478, *p* < 0.001) than in females (*β* = 0.390, 95% CI 0.374-0.407, *p* < 0.001), with a statistically significant interaction (*β* _interaction_ = 0.068, 95% CI 0.044-0.092, *p* < 0.001). Sexual assault and witnessing a sudden violent death were more strongly associated with frailty in females (*β* = 0.303, 95% CI 0.289-0.318, *p* < 0.001 and *β* = 0.218, 95% CI 0.197-0.238, *p* < 0.001, respectively) than males (*β* = 0.259, 95% CI 0.235-0.284, *p* < 0.001 and *β* = 0.153, 95% CI 0.136-0.169, *p* < 0.001, respectively), with statistically significant interactions (*β* _interaction_ = -0.040, 95% CI -0.068 to -0.011, *p* = 0.034; *β* _interaction_ = - 0.065, 95% CI -0.091 to -0.038, *p* < 0.001). Combat or war-zone exposure (*β* = -0.070, 95% CI -0.094 to -0.046, *p* < 0.001) and serious accidents (*β* = -0.038, 95% CI -0.055 to -0.021, *p* < 0.001) were associated with higher grip strength only in males. A statistically significant interaction was detected for serious accidents (*β* _interaction_ = -0.043, 95% CI -0.065 to -0.021, *p* < 0.001). Witnessing a sudden violent death was associated with higher grip strength in both sexes but the association was stronger in males (*β* _interaction_ = -0.058, 95% CI -0.078 to -0.039, *p* < 0.001). Finally, associations between life-threatening illness or physical violence and lower grip strength were stronger in males, with a statistically significant interaction for physical violence (*β* _interaction_ = 0.033, 95% CI 0.017-0.050, *p* < 0.001).

**Figure 4.**
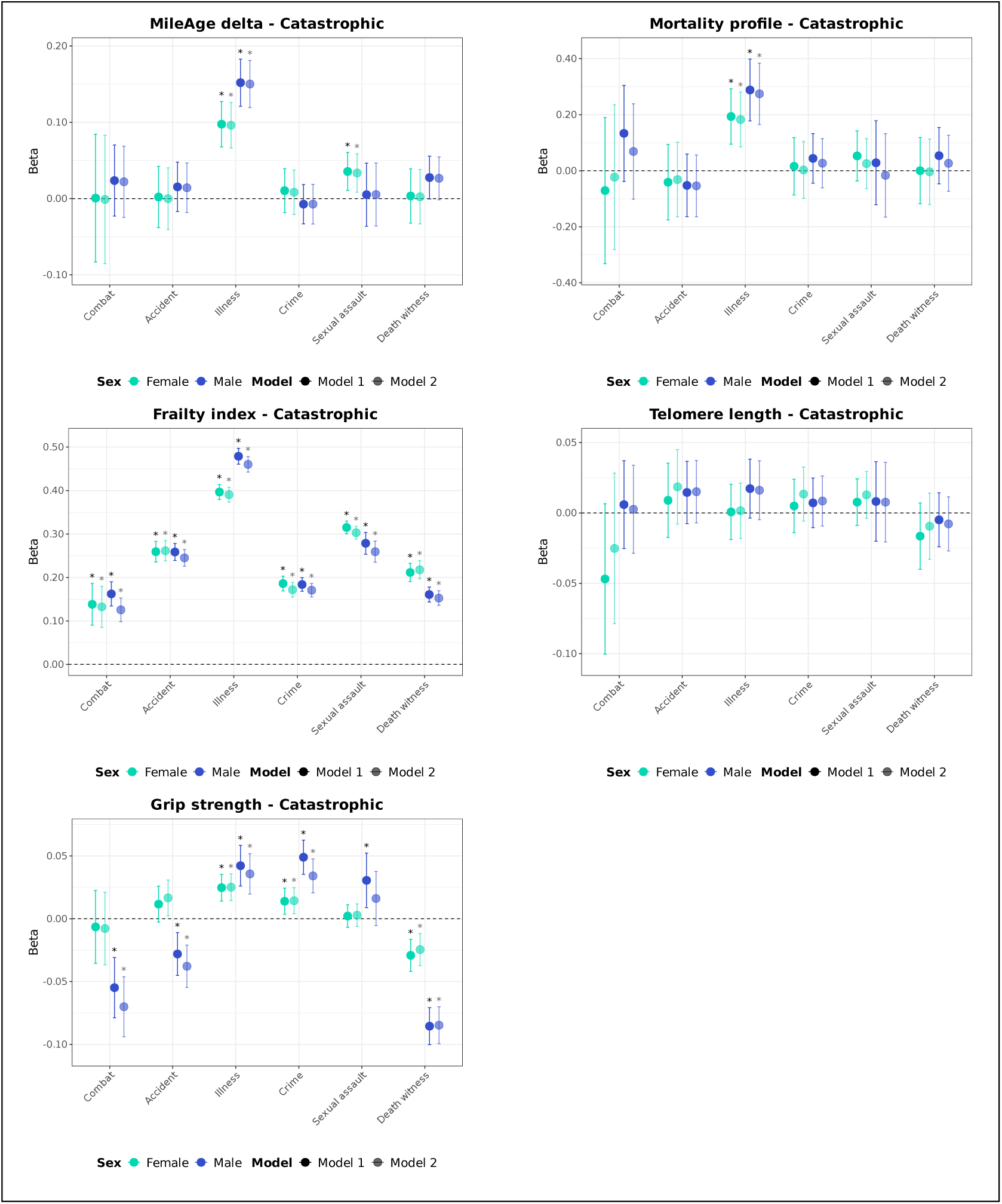
Sex-stratified associations between trauma (item-specific) and ageing markers. Sex-stratified associations between specific lifetime exposure to non-relational trauma items and ageing markers (MileAge delta, frailty index, telomere length [reverse coded] and grip strength [reverse coded]). Estimates shown are ordinary least squares regression beta coefficients and 95% confidence intervals. Model 1– adjusted for chronological age; Model 2–adjusted for chronological age, ethnicity, highest educational/professional qualification, annual gross household income and neighbourhood deprivation. Asterisks indicate statistically significant associations, after correcting *p*-values for multiple testing using the Benjamini–Hochberg procedure (across exposures, ageing markers and models, separately for each sex). Sample sizes reported in Table S7.

### Sensitivity analysis excluding sexual assault

After excluding sexual assault from the sum score, results were largely consistent with the main findings. After full adjustment, non-relational trauma was associated with higher grip strength (*β* = -0.005, 95% CI -0.009 to -0.001, *p* = 0.027) (Figure S2; Table S12). In sex-stratified analyses, trauma was associated with elevated metabolomic mortality profile scores in males after full adjustment (*β* = 0.053, 95% CI 0.010-0.096, *p* = 0.023), whereas in females this association was not statistically significant in either model (*p* ≤ 0.083) (Figure S3; Table S13). Finally, there was evidence of a statistically significant interaction between trauma and sex for grip strength (*β* _interaction_ = -0.012, 95% CI -0.020 to -0.004, *p* = 0.013) (Table S14).

## Discussion

This study demonstrates that exposure to non-relational trauma is associated with older biological ageing profiles across multiple ageing markers, including metabolomic ageing, frailty and grip strength. The strongest and most consistent association was observed for frailty across all trauma types and with evidence of a clear dose-response pattern. The use of a large, population-based sample from the UK Biobank allowed for greater statistical power to detect even moderate effect sizes compared to prior research, which has often relied on smaller samples. By examining multiple markers of biological ageing, and by distinguishing between trauma types and cumulative exposure levels, our findings provide more comprehensive insights into biological ageing as a potential mechanism underlying the long-term health consequences of trauma exposure.

Non-relational trauma was associated with a metabolite-predicted age exceeding chronological age. Metabolomic ageing clocks capture systemic metabolic dysregulation and cumulative biological stress (Huang et al., 2025). The observation that exposure to non-relational trauma was associated with an older metabolomic age suggests that such experiences may trigger or accelerate metabolic dysfunction, potentially through sustained activation of the hypothalamic-pituitary-adrenal axis, increased oxidative stress or inflammation (Heim, Ehlert, & Hellhammer, 2000; Lupien, McEwen, Gunnar, & Heim, 2009). This aligns with prior research linking trauma to dysregulation in metabolic and inflammatory pathways and underscores the potential of metabolomics as a tool to capture trauma-related changes in biological ageing (Danese & Baldwin, 2017; Mellon et al., 2018). Similarly, the metabolomic mortality profile score, a composite index trained on survival data, was elevated amongst trauma-exposed individuals. This supports the notion that trauma can have enduring consequences for health. This finding aligns with epidemiological studies identifying an increased mortality risk amongst trauma survivors and further substantiates the biological embedding of traumatic experiences (Buckley, Turiano, Sesker, Butler, & O’Súilleabháin, 2024). For both measures, life-threatening illness was the only non-relational trauma associated with advanced metabolomic ageing, suggesting that it is primarily disease-related metabolic dysregulation that is captured by these ageing profiles. Future research should examine whether perceiving an illness as life-threatening and the psychological impact of it impacts biological ageing beyond the direct physiological effects of the illness.

In contrast, associations between trauma and frailty were broad, consistent and graded, supporting frailty as the most robust measure to capture the possible impact of trauma on biological ageing. Frailty, as operationalised in this study, summarises the accumulation of age-related health deficits and functional impairments. Its sensitivity to trauma exposure highlights the broader impact that traumatic experiences may have beyond psychological outcomes such as post-traumatic stress. Our results mirror recent findings that link social stress and disadvantage with frailty in mid-to-late adulthood, suggesting that trauma may accelerate biological ageing processes via compounded physiological stress across systems (Baranyi et al., 2022). Every trauma type was associated with greater frailty, with the strongest association observed for life-threatening illness. This finding suggests that the biological ageing footprint of non-relational trauma is most pronounced when the event relates to somatic pathology.

There was no evidence that telomere length was associated with lifetime exposure to non-relational trauma. This finding is inconsistent with most studies in the field, which found greater telomere attrition in populations exposed to trauma or adversity (Aas et al., 2019; Li, He, Wang, Tang, & Chen, 2017; Oliveira et al., 2016). It is also inconsistent with our previous finding of an association between adversity experienced in childhood and shorter telomeres (Aas et al., 2025). This inconsistency with prior studies may reflect differences in sample characteristics, trauma definition and timing of exposure. For example, in our prior study (Aas et al., 2025) we examined adversity and relational trauma more broadly, whereas in the current study we focused only on non-relational trauma, which may differentially impact biological ageing.

There was also limited evidence that grip strength was associated with overall lifetime exposure to non-relational trauma. Grip strength is a strong predictor of disability and mortality and reflects the integrity of multiple systems, including neuromuscular, cardiovascular and inflammatory pathways (Gale, Martyn, Cooper, & Sayer, 2006; Rantanen et al., 1999; Xue, Walston, Fried, & Beamer, 2011). The findings reported here are largely inconsistent with our previous study generally indicating lower grip strength associated with adversity and relational trauma experienced in childhood and adulthood (Aas et al., 2025). This may be due to differences in the type and severity of exposure. Again, the present study focused on non-relational trauma only, which encompassed experiences such as military combat that may be more common among physically stronger individuals. Indeed, when examining specific types of non-relational trauma, we found that combat exposure, serious accidents or witnessing violent deaths were associated with higher grip strength, whereas life-threatening illness or being victim of a crime were associated with lower grip strength.

Importantly, a dose-response relationship was evident for frailty and, to a lesser extent, for the metabolomic clock and mortality profile score, with exposure to multiple (versus single and versus no) traumatic experiences corresponding to older biological ageing profiles. This cumulative effect is consistent with allostatic load theory, which posits that repeated or sustained stress results in wear and tear across physiological systems (Baranyi et al., 2022). The finding that individuals exposed to multiple types of non-relational trauma had older biological profiles than individuals exposed to one trauma type (and even more so than individuals not exposed to any lifetime trauma) provides empirical support for the idea of a dose-dependent impact of trauma on biological ageing.

We also identified differences in how non-relational trauma relates to biological ageing across males and females. While non-relational trauma was associated with greater frailty in both sexes, the association was stronger in females, particularly for females reporting sexual assault and/or witnessing a sudden violent death. In contrast, associations between life-threatening illness and the metabolomic clock or mortality risk score were more pronounced in males. Grip strength also showed sex-specific patterns: non-relational trauma was associated with lower grip strength in females but higher strength in males, possibly reflecting differential exposure to physically demanding trauma types, such as combat. These findings suggest that sex may moderate the biological embedding of trauma, underscoring the importance of sex-sensitive approaches in trauma research and intervention.

Our findings contribute to a growing literature on the biological embedding of trauma. While previous studies have demonstrated that early-life adversity can alter stress physiology, immune function and cellular ageing (Cunningham, Mengelkoch, Gassen, & Hill, 2022; Nusslock & Miller, 2016), fewer studies have examined adulthood trauma, particularly non-relational events (Bourassa & Sbarra, 2024). Moreover, prior studies have typically relied on single ageing biomarkers in small, clinical samples, limiting generalisability and ability to capture system-wide effects (Nelles-McGee et al., 2022). This study addresses these gaps using a large, community-based cohort and a multidimensional set of ageing markers, thereby offering a more comprehensive picture of the links between trauma and biological ageing. Importantly, future research should investigate whether trauma survivors who develop lasting mental health conditions (e.g., post-traumatic stress disorders or major depression) exhibit poorer biological outcomes compared to trauma survivors without such conditions.

These findings also have public health and clinical implications. They suggest that non-relational trauma is not only a psychological burden but also a risk factor for accelerated biological ageing, which itself is linked to premature mortality (Mutz et al., 2024). Early identification and support for individuals with trauma exposure may help mitigate some of these long-term biological consequences. Interventions aimed at reducing chronic stress, promoting physical activity, enhancing social support and addressing metabolic dysfunction may be particularly beneficial for trauma-exposed populations (Aas et al., 2021). Perhaps the need for prevention and intervention might be particularly high for those who develop mental disorders in the aftermath of trauma such as post-traumatic stress disorder and fortunately effective interventions exist (Hoppen et al., 2023). Yet, the question of whether and how trauma-related mental illnesses relate to the link between trauma and adverse physiological outcomes remains understudied for the time being.

Several limitations should be acknowledged. First, trauma exposure was retrospectively self-reported, which may introduce recall bias or underreporting. Yet, retrospective compared to prospective accounts of trauma exposure have been shown to be the clinically more useful predictors of adverse (mental) health outcomes, irrespective of recall biases (Danese & Widom, 2023). Second, the categorisation of trauma based on lifetime recall rather than timing, severity or chronicity may obscure important nuances in the exposure-response relationship. Third, the cross-sectional study design limits causal inference. Although the observed associations are biologically plausible and align with prior longitudinal research (Elliott et al., 2021; Moffitt, Belsky, Danese, Poulton, & Caspi, 2016), we cannot conclude that trauma causes accelerated biological ageing. Fourth, while we adjusted for multiple demographic and socioeconomic confounders, residual confounding is possible. For example, genetic predispositions or behavioural factors could influence both trauma exposure and biological ageing. Fifth, the UK Biobank is not fully representative of the general UK population; participants are generally healthier, wealthier and more educated, which may bias association estimates.

In conclusion, this study provides robust evidence that non-relational trauma is associated with older biological ageing profiles across multiple domains, most clearly reflected in greater frailty across all trauma types in a graded, dose-response fashion. These findings underscore the need to recognize trauma as a significant contributor to long-term physical health and biological ageing, warranting further investigation and exploring preventative and intervention strategies. Future research should focus on longitudinal designs, mechanistic pathways and modifiable resilience factors that could buffer the biological impact of trauma.

## Data Availability

The data used are available to all bona fide researchers for health-related research that is in the public interest, subject to an application process and approval criteria. Study materials are publicly available online at http://www.ukbiobank.ac.uk.

## Acknowledgements

JM is funded by the King’s Prize Fellowship. MA is funded by the MRC fellowship (#MR/W027720/1). Computational analyses were supported by King’s Computational Research, Engineering and Technology Environment (CREATE). This research has been conducted using data from UK Biobank. Data access permission has been granted under UK Biobank application 45514.

## Authorship contributions

JM and MA conceived the idea of the study. JM acquired the data and performed the statistical analysis. THH and JM wrote the manuscript. THH, NM, MA and JM interpreted the findings and revised the manuscript. All authors read and approved the final manuscript.

## Ethics

Ethical approval for the UK Biobank study has been granted by the National Information Governance Board for Health and Social Care and the NHS North West Multicentre Research Ethics Committee (11/NW/0382). No project-specific ethical approval is needed.

## Data sharing statement

The data used are available to all *bona fide* researchers for health-related research that is in the public interest, subject to an application process and approval criteria. Study materials are publicly available online at http://www.ukbiobank.ac.uk.

## Supplementary material

Supplementary information is available online.

## References

Aas, M., Elvsashagen, T., Westlye, L. T., Kaufmann, T., Athanasiu, L., Djurovic, S., . . . Andreassen, O. A. (2019). Telomere length is associated with childhood trauma in patients with severe mental disorders. Transl Psychiatry, 9(1), 97. doi:10.1038/s41398-019-0432-7

Aas, M., Hoppen, T. H., Morina, N., Zhang, S., Li, B., Mlakar, V., & Mutz, J. (2025). Adverse events in both childhood and adulthood are associated with molecular, clinical and functional markers of ageing. medRxiv, 2025.2006.2003.25328874. doi:10.1101/2025.06.03.25328874

Aas, M., Ueland, T., Mørch, R. H., Laskemoen, J. F., Lunding, S. H., Reponen, E. J., . . . Andreassen, O. A. (2021). Physical activity and childhood trauma experiences in patients with schizophrenia or bipolar disorders. The World Journal of Biological Psychiatry, 22(8), 637–645. doi:10.1080/15622975.2021.1907707

Austad, S. N., & Fischer, K. E. (2016). Sex Differences in Lifespan. Cell Metabolism, 23(6), 1022–1033. doi:10.1016/j.cmet.2016.05.019

Baranyi, G., Welstead, M., Corley, J., Deary, I. J., Muniz-Terrera, G., Redmond, P., . . . Pearce, J. (2022). Association of Life-Course Neighborhood Deprivation With Frailty and Frailty Progression From Ages 70 to 82 Years in the Lothian Birth Cohort 1936. American Journal of Epidemiology, 191(11), 1856–1866. doi:10.1093/aje/kwac134

Bourassa, K. J., & Sbarra, D. A. (2024). Trauma, adversity, and biological aging: behavioral mechanisms relevant to treatment and theory. Translational Psychiatry, 14(1), 285. doi:10.1038/s41398-024-03004-9

Buckley, L., Turiano, N., Sesker, A., Butler, M., & O’Súilleabháin, P. S. (2024). Lifetime trauma and mortality risk: A systematic review. Health Psychology, 43(4), 280–288. doi:10.1037/hea0001343

Codd, V., Denniff, M., Swinfield, C., Warner, S. C., Papakonstantinou, M., Sheth, S., . . . Samani, N. J. (2022). Measurement and initial characterization of leukocyte telomere length in 474,074 participants in UK Biobank. Nat Aging, 2(2), 170–179. doi:10.1038/s43587-021-00166-9

Cunningham, K., Mengelkoch, S., Gassen, J., & Hill, S. E. (2022). Early life adversity, inflammation, and immune function: An initial test of adaptive response models of immunological programming. Development and Psychopathology, 34(2), 539–555. doi:10.1017/S095457942100170X

Danese, A., & Baldwin, J. R. (2017). Hidden Wounds? Inflammatory Links Between Childhood Trauma and Psychopathology. Annual Review of Psychology, 68(Volume 68, 2017), 517–544. 10.1146/annurev-psych-010416-044208

Danese, A., & Widom, C. S. (2023). Associations Between Objective and Subjective Experiences of Childhood Maltreatment and the Course of Emotional Disorders in Adulthood. JAMA Psychiatry, 80(10), 1009–1016. doi:10.1001/jamapsychiatry.2023.2140

Davis, K. A. S., Coleman, J. R. I., Adams, M., Allen, N., Breen, G., Cullen, B., . . . Hotopf, M. (2020). Mental health in UK Biobank -development, implementation and results from an online questionnaire completed by 157 366 participants: a reanalysis. BJPsych Open, 6(2), e18. doi:10.1192/bjo.2019.100

Duchowny, K. A., Marcinek, D. J., Mau, T., Diaz-Ramierz, L. G., Lui, L.-Y., Toledo, F. G. S., . . . Molina, A. J. A. (2024). Childhood adverse life events and skeletal muscle mitochondrial function. Science Advances, 10(10), eadj6411. doi:doi:10.1126/sciadv.adj6411

Elliott, M. L., Caspi, A., Houts, R. M., Ambler, A., Broadbent, J. M., Hancox, R. J., . . . Moffitt, T. E. (2021). Disparities in the pace of biological aging among midlife adults of the same chronological age have implications for future frailty risk and policy. Nature Aging, 1(3), 295–308. doi:10.1038/s43587-021-00044-4

Epel, E. S., Blackburn, E. H., Lin, J., Dhabhar, F. S., Adler, N. E., Morrow, J. D., & Cawthon, R. M. (2004). Accelerated telomere shortening in response to life stress. Proceedings of the National Academy of Sciences, 101(49), 17312–17315. doi:doi:10.1073/pnas.0407162101

Fiehn, O. (2002). Metabolomics — the link between genotypes and phenotypes. In C. Town (Ed.), Functional Genomics (pp. 155–171). Dordrecht: Springer Netherlands.

Frissa, S., Hatch, S. L., Fear, N. T., Dorrington, S., Goodwin, L., & Hotopf, M. (2016). Challenges in the retrospective assessment of trauma: comparing a checklist approach to a single item trauma experience screening question. BMC Psychiatry, 16, 20. doi:10.1186/s12888-016-0720-1

Furrer, R., & Handschin, C. (2025). Biomarkers of aging: functional aspects still trump molecular parameters. npj Aging, 11(1), 15. doi:10.1038/s41514-025-00207-2

Gale, C. R., Martyn, C. N., Cooper, C., & Sayer, A. A. (2006). Grip strength, body composition, and mortality. International Journal of Epidemiology, 36(1), 228–235. doi:10.1093/ije/dyl224

Grummitt, L. R., Kreski, N. T., Kim, S. G., Platt, J., Keyes, K. M., & McLaughlin, K. A. (2021). Association of Childhood Adversity With Morbidity and Mortality in US Adults: A Systematic Review. JAMA Pediatrics, 175(12), 1269–1278. doi:10.1001/jamapediatrics.2021.2320

Hägg, S., & Jylhävä, J. (2021). Sex differences in biological aging with a focus on human studies. eLife, 10, e63425. doi:10.7554/eLife.63425

Heim, C., Ehlert, U., & Hellhammer, D. H. (2000). The potential role of hypocortisolism in the pathophysiology of stress-related bodily disorders. Psychoneuroendocrinology, 25(1), 1–35. 10.1016/S0306-4530(99)00035-9

Hoppen, T. H., Jehn, M., Holling, H., Mutz, J., Kip, A., & Morina, N. (2023). The efficacy and acceptability of psychological interventions for adult PTSD: A network and pairwise meta-analysis of randomized controlled trials. J Consult Clin Psychol, 91(8), 445–461. doi:10.1037/ccp0000809

Hoppen, T. H., Priebe, S., Vetter, I., & Morina, N. (2021). Global burden of post-traumatic stress disorder and major depression in countries affected by war between 1989 and 2019: a systematic review and meta-analysis. BMJ Global Health, 6(7), e006303. doi:10.1136/bmjgh-2021-006303

Huang, H., Chen, Y., Xu, W., Cao, L., Qian, K., Bischof, E., . . . Pu, J. (2025). Decoding aging clocks: New insights from metabolomics. Cell Metabolism, 37(1), 34–58. doi:10.1016/j.cmet.2024.11.007

Kessler, R. C., Aguilar-Gaxiola, S., Alonso, J., Benjet, C., Bromet, E. J., Cardoso, G., . . . Koenen, K. C. (2017). Trauma and PTSD in the WHO World Mental Health Surveys. European Journal of Psychotraumatology, 8(sup5), 1353383. doi:10.1080/20008198.2017.1353383

Lai, T. P., Wright, W. E., & Shay, J. W. (2018). Comparison of telomere length measurement methods. Philos Trans R Soc Lond B Biol Sci, 373(1741). doi:10.1098/rstb.2016.0451

Lee, S. H., Shin, J., Um, S., Shin, H. R., Kim, Y. S., & Choi, J. K. (2023). Perceived Stress and Frailty in Older Adults. Ann Geriatr Med Res, 27(4), 310–314. doi:10.4235/agmr.23.0132

Li, Z., He, Y., Wang, D., Tang, J., & Chen, X. (2017). Association between childhood trauma and accelerated telomere erosion in adulthood: A meta-analytic study. Journal of Psychiatric Research, 93, 64–71. 10.1016/j.jpsychires.2017.06.002

Lupien, S. J., McEwen, B. S., Gunnar, M. R., & Heim, C. (2009). Effects of stress throughout the lifespan on the brain, behaviour and cognition. Nature Reviews Neuroscience, 10(6), 434–445. doi:10.1038/nrn2639

Mellon, S. H., Gautam, A., Hammamieh, R., Jett, M., & Wolkowitz, O. M. (2018). Metabolism, Metabolomics, and Inflammation in Posttraumatic Stress Disorder. Biological Psychiatry, 83(10), 866–875. 10.1016/j.biopsych.2018.02.007

Moffitt, T. E., Belsky, D. W., Danese, A., Poulton, R., & Caspi, A. (2016). The Longitudinal Study of Aging in Human Young Adults: Knowledge Gaps and Research Agenda. The Journals of Gerontology: Series A, 72(2), 210–215. doi:10.1093/gerona/glw191

Mutz, J., Choudhury, U., Zhao, J., & Dregan, A. (2022). Frailty in individuals with depression, bipolar disorder and anxiety disorders: longitudinal analyses of all-cause mortality. BMC Med, 20(1), 274. doi:10.1186/s12916-022-02474-2

Mutz, J., Iniesta, R., & Lewis, C. (2024). Metabolomic Age (MileAge) predicts health and lifespan: a comparison of multiple machine learning algorithms. Science Advances, *In Press*. doi:10.1126/sciadv.adp3743

Mutz, J., & Lewis, C. (2021). Lifetime depression and age-related changes in body composition, cardiovascular function, grip strength and lung function: sex-specific analyses in the UK Biobank. Aging, 13(13), 17038–17079. doi:10.18632/aging.203275

Nelles-McGee, T., Khoury, J., Kenny, M., Joshi, D., & Gonzalez, A. (2022). Biological embedding of child maltreatment: A systematic review of biomarkers and resilience in children and youth. Psychol Trauma, 14(S1), S50–s62. doi:10.1037/tra0001162

Nusslock, R., & Miller, G. E. (2016). Early-Life Adversity and Physical and Emotional Health Across the Lifespan: A Neuroimmune Network Hypothesis. Biological Psychiatry, 80(1), 23–32. 10.1016/j.biopsych.2015.05.017

Oliveira, B. S., Zunzunegui, M. V., Quinlan, J., Fahmi, H., Tu, M. T., & Guerra, R. O. (2016). Systematic review of the association between chronic social stress and telomere length: A life course perspective. Ageing Research Reviews, 26, 37–52. 10.1016/j.arr.2015.12.006

Pickhardt, P. J., Kattan, M. W., Lee, M. H., Pooler, B. D., Pyrros, A., Liu, D., . . . Garrett, J. W. (2025). Biological age model using explainable automated CT-based cardiometabolic biomarkers for phenotypic prediction of longevity. Nature Communications, 16(1), 1432. doi:10.1038/s41467-025-56741-w

Rantanen, T., Guralnik, J. M., Foley, D., Masaki, K., Leveille, S., Curb, J. D., & White, L. (1999). Midlife Hand Grip Strength as a Predictor of Old Age Disability. JAMA, 281(6), 558–560. doi:10.1001/jama.281.6.558

Ritchie, S., Surendran, P., Karthikeyan, S., Lambert, S., Bolton, T., Pennells, L., . . . Inouye, M. (2023). Quality control and removal of technical variation of NMR metabolic biomarker data in ∼120,000 UK Biobank participants. Sci Data, 10(1), 64. doi:10.1038/s41597-023-01949-y

Teschendorff, A. E., & Horvath, S. (2025). Epigenetic ageing clocks: statistical methods and emerging computational challenges. Nature Reviews Genetics, 26(5), 350–368. doi:10.1038/s41576-024-00807-w

Tolin, D. F., & Foa, E. B. (2006). Sex differences in trauma and posttraumatic stress disorder: a quantitative review of 25 years of research. Psychol Bull, 132(6), 959–992. doi:10.1037/0033-2909.132.6.959

Williams, D. M., Jylhävä, J., Pedersen, N. L., & Hägg, S. (2019). A Frailty Index for UK Biobank Participants. J Gerontol A Biol Sci Med Sci, 74(4), 582–587. doi:10.1093/gerona/gly094

Würtz, P., Kangas, A. J., Soininen, P., Lawlor, D. A., Davey Smith, G., & Ala-Korpela, M. (2017). Quantitative Serum Nuclear Magnetic Resonance Metabolomics in Large-Scale Epidemiology: A Primer on -Omic Technologies. American Journal of Epidemiology, 186(9), 1084–1096. doi:10.1093/aje/kwx016

Xue, Q.-L., Walston, J. D., Fried, L. P., & Beamer, B. A. (2011). Prediction of Risk of Falling, Physical Disability, and Frailty by Rate of Decline in Grip Strength: The Women’s Health and Aging Study. Archives of Internal Medicine, 171(12), 1119–1121. doi:10.1001/archinternmed.2011.252

Zhang, S., Wang, Z., Wang, Y., Zhu, Y., Zhou, Q., Jian, X., . . . Li, B. (2024). A metabolomic profile of biological aging in 250,341 individuals from the UK Biobank. Nature Communications, 15(1), 8081. doi:10.1038/s41467-024-52310-9

