## Supplementary material for "Lifetime non-relational traumatic experiences and biological ageing"

This material accompanies the article

### 1. Sample characteristics

**Table S1.** Sample characteristics stratified by non-relational trauma

| | Non-relational trauma | | |
|---|---|---|---|
| | No (*N*=75,806) | Yes (*N*=77,057) | Full sample (*N*=152,863) |
| MileAge delta, mean (SD)[1] | -0.10 (3.73) | 0.03 (3.79) | -0.04 (3.76) |
| Mortality profile, mean (SD)[1] | -45.15 (0.51) | -45.08 (0.54) | -45.11 (0.52) |
| Frailty index, mean (SD)[1] | 0.10 (0.06) | 0.12 (0.07) | 0.11 (0.07) |
| T/S ratio, mean (SD)[1] | 0.06 (0.99) | 0.05 (0.99) | 0.05 (0.99) |
| Grip strength, mean (SD)[1] | 31.62 (10.74) | 33.21 (11.24) | 32.42 (11.03) |
| Age, mean (SD) | 56.59 (7.72) | 56.22 (7.74) | 56.40 (7.73) |
| Sex | | | |
| Female | 46,361 (61.2%) | 39,950 (51.8%) | 86,311 (56.5%) |
| Male | 29,445 (38.8%) | 37,107 (48.2%) | 66,552 (43.5%) |
| Ethnicity | | | |
| White | 73,662 (97.2%) | 74,316 (96.4%) | 147,978 (96.8%) |
| Mixed | 278 (0.4%) | 525 (0.7%) | 803 (0.5%) |
| Black | 504 (0.7%) | 591 (0.8%) | 1095 (0.7%) |
| Asian | 629 (0.8%) | 654 (0.8%) | 1283 (0.8%) |
| Chinese | 200 (0.3%) | 138 (0.2%) | 338 (0.2%) |
| Other | 335 (0.4%) | 511 (0.7%) | 846 (0.6%) |
| Missing[2] | 198 (0.3%) | 322 (0.4%) | 520 (0.3%) |
| Highest qualification | | | |
| None | 5,848 (7.7%) | 4,630 (6.0%) | 10,478 (6.9%) |
| O levels/GCSEs/CSEs | 18,718 (24.7%) | 17,067 (22.1%) | 35,785 (23.4%) |
| A levels/NVQ/HND/HNC[3] | 17,963 (23.7%) | 17,915 (23.2%) | 35,878 (23.5%) |
| Degree | 32,570 (43.0%) | 36,717 (47.6%) | 69,287 (45.3%) |
| Missing[2] | 707 (0.9%) | 728 (0.9%) | 1,435 (0.9%) |
| Household income[4] | | | |
| Very low | 8,978 (11.8%) | 9,897 (12.8%) | 18,875 (12.3%) |
| Low | 16,196 (21.4%) | 15,932 (20.7%) | 32,128 (21.0%) |
| Medium | 19,724 (26.0%) | 20,178 (26.2%) | 39,902 (26.1%) |
| High | 17,542 (23.1%) | 18,311 (23.8%) | 35,853 (23.5%) |
| Very high | 5,134 (6.8%) | 5,876 (7.6%) | 11,010 (7.2%) |
| Missing[2] | 8,232 (10.9%) | 6,863 (8.9%) | 15,095 (9.9%) |
| Townsend deprivation | | | |
| Q1 | 18,709 (24.7%) | 15,859 (20.6%) | 34,568 (22.6%) |
| Q2 | 17,350 (22.9%) | 15,491 (20.1%) | 32,841 (21.5%) |
| Q3 | 15,975 (21.1%) | 15,448 (20.0%) | 31,423 (20.6%) |
| Q4 | 14,082 (18.6%) | 15,932 (20.7%) | 30,014 (19.6%) |
| Q5 | 9,599 (12.7%) | 14,224 (18.5%) | 23,823 (15.6%) |
| Missing[2] | 91 (0.1%) | 103 (0.1%) | 194 (0.1%) |

*Note:* Numbers shown are counts and percentages unless indicated otherwise. SD = standard deviation. GCSEs = general certificate of secondary education; CSE = certificate of secondary education; NVQ = national vocational qualification; HND = higher national diploma; HNC = higher national certificate. [1] Missing data for health and ageing markers: *n* = 69,042 (MileAge delta), 4,252 (mortality profile), 152,689 (frailty index), 144,288 (T/S ratio) and 152,366 (grip strength). [2] Missing data may also include "do not know" or "prefer not to answer". [3] Also includes 'other professional qualifications'. [4] Annual household income groups: very low (<£18,000), low (£18,000–£30,999), middle (£31,000–£51,999), high (£52,000–£100,000) and very high (>£100,000).

#### 2. Analytical sample sizes

**Table S2.** Non-relational trauma analytical sample sizes

| Non-relational trauma | MileAge delta | Mortality profile | Frailty index | Telomere length | Grip strength |
|---|---|---|---|---|---|
| No | 34,695 (50.3%) | 2,174 (51.1%) | 75,730 (49.6%) | 71,618 (49.6%) | 75,578 (49.6%) |
| Yes | 34,347 (49.7%) | 2,078 (48.9%) | 76,959 (50.4%) | 72,670 (50.4%) | 76,788 (50.4%) |
| 0 | 34,695 (51.6%) | 2,174 (52.5%) | 75,730 (50.9%) | 71,618 (51.0%) | 75,578 (50.9%) |
| 1 | 21,003 (31.2%) | 1,252 (30.2%) | 46,639 (31.4%) | 44,051 (31.4%) | 46,541 (31.4%) |
| 2 | 8,237 (12.2%) | 535 (12.9%) | 18,695 (12.6%) | 17,629 (12.5%) | 18,651 (12.6%) |
| 3 | 2,508 (3.7%) | 135 (3.3%) | 5,772 (3.9%) | 5,466 (3.9%) | 5,745 (3.9%) |
| 4 | 652 (1.0%) | 38 (0.9%) | 1,520 (1.0%) | 1,415 (1.0%) | 1,517 (1.0%) |
| 5/6 | 149 (0.2%) | 10 (0.2%) | 340 (0.2%) | 326 (0.2%) | 340 (0.2%) |
| None | 34,695 (51.6%) | 2,174 (52.5%) | 75,730 (50.9%) | 71,618 (51.0%) | 75,578 (50.9%) |
| One | 21,003 (31.2%) | 1,252 (30.2%) | 46,639 (31.4%) | 44,051 (31.4%) | 46,541 (31.4%) |
| Multiple | 11,546 (17.2%) | 718 (17.3%) | 26,327 (17.7%) | 24,836 (17.7%) | 26,253 (17.7%) |
| Sum score[1] | 0.72 (0.91) | 0.71 (0.91) | 0.73 (0.92) | 0.73 (0.92) | 0.73 (0.92) |
| Sum score (sens.)[1] | 0.57 (0.81) | 0.58 (0.81) | 0.59 (0.82) | 0.59 (0.82) | 0.59 (0.82) |
| Specific items | | | | | |
| Combat | 68,522 (96.7%) | 4,215 (96.8%) | 151,292 (96.5%) | 142,933 (96.5%) | 150,966 (96.5%) |
| Yes | 2,371 (3.3%) | 141 (3.2%) | 5,508 (3.5%) | 5,231 (3.5%) | 5,503 (3.5%) |
| Accident | 64,088 (90.6%) | 3,913 (90.2%) | 141,530 (90.5%) | 133,718 (90.5%) | 141,232 (90.5%) |
| Yes | 6,635 (9.4%) | 427 (9.8%) | 14,903 (9.5%) | 14,081 (9.5%) | 14,866 (9.5%) |
| Illness | 59,569 (86.0%) | 3,659 (86.0%) | 130,903 (85.5%) | 123,786 (85.6%) | 130,635 (85.5%) |
| Yes | 9,702 (14.0%) | 597 (14.0%) | 22,170 (14.5%) | 20,835 (14.4%) | 22,116 (14.5%) |
| Crime | 57,535 (81.5%) | 3,540 (81.5%) | 126,885 (81.3%) | 119,869 (81.3%) | 126,622 (81.3%) |
| Yes | 13,033 (18.5%) | 804 (18.5%) | 29,147 (18.7%) | 27,575 (18.7%) | 29,078 (18.7%) |
| Sexual assault | 59,825 (85.4%) | 3,745 (87.0%) | 131,850 (85.1%) | 124,650 (85.1%) | 131,569 (85.1%) |
| Yes | 10,233 (14.6%) | 562 (13.0%) | 23,094 (14.9%) | 21,772 (14.9%) | 23,043 (14.9%) |
| Death witness | 61,401 (87.1%) | 3,788 (87.3%) | 135,634 (87.0%) | 128,160 (87.0%) | 135,363 (87.0%) |
| Yes | 9,113 (12.9%) | 549 (12.7%) | 20,335 (13.0%) | 19,218 (13.0%) | 20,276 (13.0%) |

*Note:* Numbers shown are counts and percentages unless indicated otherwise. [1] mean and standard deviation.

#### 3. Multiple non-relational traumas

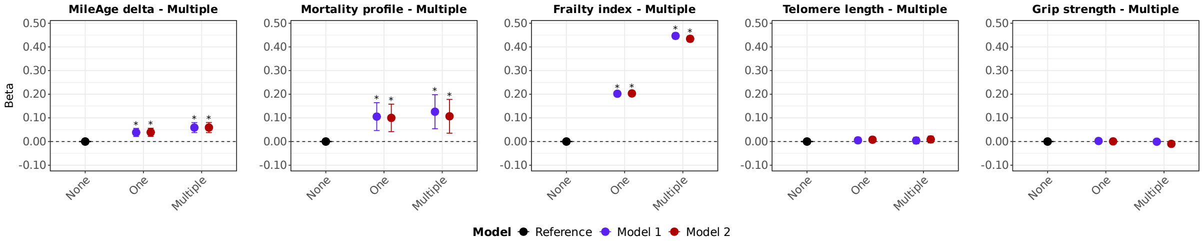

**Figure S1. Multiple traumas and ageing markers.** Associations between lifetime exposure to multiple non-relational traumas and ageing markers (MileAge delta, frailty index, telomere length [reverse coded] and grip strength [reverse coded]). Estimates shown are ordinary least squares regression beta coefficients and 95% confidence intervals. Model 1–adjusted for chronological age and sex; Model 2–adjusted for chronological age, sex, ethnicity, education, income and neighbourhood deprivation. Asterisks indicate statistically significant associations, after correcting *p*-values for multiple testing using the Benjamini–Hochberg procedure (across exposure levels and models, separately for each ageing marker. Sample sizes reported in Tables S2.

**Table S3.** Associations between exposure to multiple non-relational traumas and ageing markers

| | Model 1 (adj. age and sex) | | | | Model 2 (full adjustment) | | | |
|---|---|---|---|---|---|---|---|---|
| Trauma exposure | *β* | 95% CI | | *p* | *β* | 95% CI | | *p* |
| MileAge delta | | | | | | | | |
| None (Reference) | | | | | | | | |
| One | 0.038 | 0.021 | 0.056 | <0.001 | 0.039 | 0.022 | 0.056 | <0.001 |
| Multiple | 0.059 | 0.038 | 0.080 | <0.001 | 0.059 | 0.038 | 0.080 | <0.001 |
| Metabolomic profile | | | | | | | | |
| None (Reference) | | | | | | | | |
| One | 0.105 | 0.046 | 0.164 | 0.001 | 0.100 | 0.042 | 0.158 | 0.001 |
| Multiple | 0.126 | 0.054 | 0.198 | 0.001 | 0.107 | 0.035 | 0.178 | 0.003 |
| Frailty index | | | | | | | | |
| None (Reference) | | | | | | | | |
| One | 0.202 | 0.192 | 0.212 | <0.001 | 0.203 | 0.193 | 0.213 | <0.001 |
| Multiple | 0.447 | 0.434 | 0.459 | <0.001 | 0.434 | 0.422 | 0.446 | <0.001 |
| Telomere length | | | | | | | | |
| None (Reference) | | | | | | | | |
| One | 0.005 | -0.006 | 0.017 | 0.513 | 0.008 | -0.004 | 0.019 | 0.433 |
| Multiple | 0.004 | -0.010 | 0.018 | 0.546 | 0.009 | -0.005 | 0.023 | 0.433 |
| Grip strength | | | | | | | | |
| None (Reference) | | | | | | | | |
| One | 0.002 | -0.005 | 0.010 | 0.886 | 0.001 | -0.007 | 0.008 | 0.886 |
| Multiple | -0.001 | -0.010 | 0.008 | 0.886 | -0.010 | -0.019 | -0.001 | 0.139 |

*Note:* CI = confidence interval. Model 1–adjusted for chronological age and sex; Model 2–adjusted for chronological age, sex, ethnicity, highest educational/professional qualification, annual gross household income and Townsend deprivation index. *P*-values shown are corrected for multiple testing using the Benjamini–Hochberg procedure (across exposure levels and models, separately for each ageing marker). Sample sizes reported in Table S2.

#### 4. Non-relational traumas (ordinal)

**Table S4.** Associations between non-relational trauma exposures and ageing markers (ordinal)

| | Model 1 (adj. age and sex) | | | | Model 2 (full adjustment) | | | |
|---|---|---|---|---|---|---|---|---|
| Trauma exposure | *β* | 95% CI | | *p* | *β* | 95% CI | | *p* |
| MileAge delta | | | | | | | | |
| None (Reference) | | | | | | | | |
| 1 | 0.038 | 0.021 | 0.056 | <0.001 | 0.039 | 0.022 | 0.056 | <0.001 |
| 2 | 0.060 | 0.036 | 0.084 | <0.001 | 0.060 | 0.036 | 0.084 | <0.001 |
| 3 | 0.042 | 0.001 | 0.082 | 0.056 | 0.042 | 0.001 | 0.082 | 0.056 |
| 4 | 0.118 | 0.041 | 0.195 | 0.005 | 0.117 | 0.040 | 0.194 | 0.005 |
| 5/6 | 0.059 | -0.101 | 0.220 | 0.482 | 0.058 | -0.103 | 0.218 | 0.482 |
| Metabolomic profile | | | | | | | | |
| None (Reference) | | | | | | | | |
| 1 | 0.105 | 0.047 | 0.164 | 0.003 | 0.100 | 0.042 | 0.158 | 0.003 |
| 2 | 0.140 | 0.059 | 0.220 | 0.003 | 0.127 | 0.047 | 0.206 | 0.004 |
| 3 | 0.056 | -0.091 | 0.204 | 0.506 | 0.014 | -0.132 | 0.160 | 0.854 |
| 4 | 0.291 | 0.020 | 0.562 | 0.071 | 0.256 | -0.012 | 0.524 | 0.102 |
| 5/6 | -0.296 | -0.821 | 0.229 | 0.336 | -0.310 | -0.829 | 0.209 | 0.336 |
| Frailty index | | | | | | | | |
| None (Reference) | | | | | | | | |
| 1 | 0.202 | 0.192 | 0.212 | <0.001 | 0.204 | 0.194 | 0.214 | <0.001 |
| 2 | 0.378 | 0.364 | 0.392 | <0.001 | 0.371 | 0.357 | 0.384 | <0.001 |
| 3 | 0.563 | 0.540 | 0.586 | <0.001 | 0.545 | 0.522 | 0.568 | <0.001 |
| 4 | 0.719 | 0.675 | 0.763 | <0.001 | 0.686 | 0.642 | 0.729 | <0.001 |
| 5/6 | 1.071 | 0.978 | 1.163 | <0.001 | 1.024 | 0.933 | 1.114 | <0.001 |
| Telomere length | | | | | | | | |
| None (Reference) | | | | | | | | |
| 1 | 0.005 | -0.006 | 0.017 | 0.629 | 0.008 | -0.004 | 0.019 | 0.417 |
| 2 | 0.000 | -0.016 | 0.016 | >0.999 | 0.005 | -0.011 | 0.021 | 0.699 |
| 3 | 0.007 | -0.020 | 0.033 | 0.699 | 0.011 | -0.016 | 0.037 | 0.629 |
| 4 | 0.033 | -0.018 | 0.084 | 0.417 | 0.037 | -0.014 | 0.089 | 0.417 |
| 5/6 | 0.081 | -0.024 | 0.187 | 0.417 | 0.094 | -0.012 | 0.199 | 0.417 |
| Grip strength | | | | | | | | |
| None (Reference) | | | | | | | | |
| 1 | 0.002 | -0.005 | 0.010 | 0.913 | 0.001 | -0.007 | 0.008 | 0.969 |
| 2 | 0.000 | -0.011 | 0.010 | 0.969 | -0.007 | -0.018 | 0.003 | 0.573 |
| 3 | -0.010 | -0.028 | 0.007 | 0.609 | -0.022 | -0.040 | -0.005 | 0.114 |
| 4 | 0.023 | -0.010 | 0.056 | 0.573 | 0.003 | -0.030 | 0.036 | 0.969 |
| 5/6 | 0.033 | -0.036 | 0.102 | 0.706 | 0.006 | -0.063 | 0.074 | 0.969 |

*Note:* CI = confidence interval. Model 1–adjusted for chronological age and sex; Model 2–adjusted for chronological age, sex, ethnicity, highest educational/professional qualification, annual gross household income and Townsend deprivation index. *P*-values shown are corrected for multiple testing using the Benjamini–Hochberg procedure (across exposure levels and models, separately for each ageing marker). Sample sizes reported in Table S2.

#### 5. Non-relational traumas (item-specific)

**Table S5** Associations between non-relational trauma items and ageing markers

| | Model 1 (adj. age and sex) | | | | Model 2 (full adjustment) | | | |
|---|---|---|---|---|---|---|---|---|
| Trauma exposure | *β* | 95% CI | | *p* | *β* | 95% CI | | *p* |
| MileAge delta | | | | | | | | |
| Combat | 0.010 | -0.031 | 0.051 | 0.654 | 0.009 | -0.032 | 0.051 | 0.654 |
| Accident | 0.012 | -0.013 | 0.038 | 0.461 | 0.011 | -0.014 | 0.037 | 0.468 |
| Illness | 0.118 | 0.096 | 0.139 | <0.001 | 0.117 | 0.095 | 0.138 | <0.001 |
| Crime | 0.013 | -0.007 | 0.032 | 0.348 | 0.012 | -0.008 | 0.031 | 0.357 |
| Sexual assault | 0.017 | -0.004 | 0.038 | 0.304 | 0.016 | -0.005 | 0.038 | 0.304 |
| Death witness | 0.017 | -0.006 | 0.039 | 0.304 | 0.016 | -0.006 | 0.038 | 0.304 |
| Metabolomic profile | | | | | | | | |
| Combat | 0.076 | -0.067 | 0.220 | 0.595 | 0.058 | -0.084 | 0.200 | 0.599 |
| Accident | -0.051 | -0.138 | 0.035 | 0.595 | -0.046 | -0.131 | 0.039 | 0.595 |
| Illness | 0.239 | 0.165 | 0.313 | <0.001 | 0.224 | 0.151 | 0.297 | <0.001 |
| Crime | 0.026 | -0.041 | 0.093 | 0.599 | 0.012 | -0.054 | 0.078 | 0.725 |
| Sexual assault | 0.051 | -0.025 | 0.128 | 0.595 | 0.025 | -0.052 | 0.101 | 0.633 |
| Death witness | 0.031 | -0.046 | 0.107 | 0.599 | 0.019 | -0.057 | 0.095 | 0.679 |
| Frailty index | | | | | | | | |
| Combat | 0.156 | 0.132 | 0.180 | <0.001 | 0.129 | 0.105 | 0.152 | <0.001 |
| Accident | 0.259 | 0.244 | 0.274 | <0.001 | 0.252 | 0.237 | 0.267 | <0.001 |
| Illness | 0.434 | 0.421 | 0.446 | <0.001 | 0.422 | 0.410 | 0.435 | <0.001 |
| Crime | 0.185 | 0.173 | 0.196 | <0.001 | 0.170 | 0.159 | 0.182 | <0.001 |
| Sexual assault | 0.305 | 0.292 | 0.317 | <0.001 | 0.291 | 0.279 | 0.304 | <0.001 |
| Death witness | 0.181 | 0.168 | 0.194 | <0.001 | 0.180 | 0.167 | 0.193 | <0.001 |
| Telomere length | | | | | | | | |
| Combat | -0.006 | -0.033 | 0.021 | 0.783 | -0.004 | -0.031 | 0.024 | 0.800 |
| Accident | 0.011 | -0.006 | 0.028 | 0.371 | 0.015 | -0.002 | 0.032 | 0.371 |
| Illness | 0.010 | -0.004 | 0.025 | 0.371 | 0.010 | -0.004 | 0.024 | 0.371 |
| Crime | 0.002 | -0.011 | 0.015 | 0.796 | 0.006 | -0.007 | 0.019 | 0.440 |
| Sexual assault | 0.011 | -0.003 | 0.025 | 0.371 | 0.014 | 0.000 | 0.029 | 0.371 |
| Death witness | -0.009 | -0.024 | 0.006 | 0.390 | -0.008 | -0.023 | 0.006 | 0.400 |
| Grip strength | | | | | | | | |
| Combat | -0.042 | -0.060 | -0.025 | <0.001 | -0.057 | -0.075 | -0.040 | <0.001 |
| Accident | -0.012 | -0.023 | -0.001 | 0.044 | -0.016 | -0.027 | -0.006 | 0.004 |
| Illness | 0.033 | 0.024 | 0.042 | <0.001 | 0.030 | 0.021 | 0.039 | <0.001 |
| Crime | 0.032 | 0.024 | 0.040 | <0.001 | 0.023 | 0.015 | 0.032 | <0.001 |
| Sexual assault | 0.010 | 0.000 | 0.019 | 0.044 | 0.004 | -0.006 | 0.013 | 0.456 |
| Death witness | -0.063 | -0.072 | -0.053 | <0.001 | -0.063 | -0.073 | -0.053 | <0.001 |

*Note:* CI = confidence interval. Model 1–adjusted for chronological age and sex; Model 2–adjusted for chronological age, sex, ethnicity, highest educational/professional qualification, annual gross household income and Townsend deprivation index. *P*-values shown are corrected for multiple testing using the Benjamini–Hochberg procedure (across trauma items and models, separately for each ageing marker). Sample sizes reported in Table S2.

#### 6. Sample characteristics stratified by sex

**Table S6.** Sample characteristics stratified by non-relational trauma and sex

| | Non-relational trauma | | | | |
|---|---|---|---|---|---|
| | Female | | Male | | |
| | No (*N*=46,361) | Yes (*N*=39,950) | No (*N*=29,445) | Yes (*N*=37,107) | Full sample (*N*=152,863) |
| MileAge delta, mean (SD)[1] | 0.08 (3.77) | 0.25 (3.84) | -0.38 (3.67) | -0.20 (3.73) | -0.04 (3.76) |
| Mortality profile, mean (SD)[1] | -45.19 (0.50) | -45.14 (0.53) | -45.07 (0.50) | -45.01 (0.53) | -45.11 (0.52) |
| Frailty index, mean (SD)[1] | 0.10 (0.06) | 0.13 (0.07) | 0.10 (0.06) | 0.12 (0.07) | 0.11 (0.07) |
| T/S ratio, mean (SD)[1] | 0.13 (0.99) | 0.15 (0.98) | -0.06 (0.97) | -0.06 (0.99) | 0.05 (0.99) |
| Grip strength, mean (SD)[1] | 25.40 (6.37) | 25.45 (6.60) | 41.40 (8.75) | 41.55 (9.02) | 32.42 (11.03) |
| Age, mean (SD) | 56.16 (7.67) | 55.65 (7.61) | 57.27 (7.75) | 56.83 (7.82) | 56.40 (7.73) |
| Ethnicity | | | | | |
| White | 45,143 (97.4%) | 38,453 (96.3%) | 28,519 (96.9%) | 35,863 (96.6%) | 147,978 (96.8%) |
| Mixed | 192 (0.4%) | 324 (0.8%) | 86 (0.3%) | 201 (0.5%) | 803 (0.5%) |
| Black | 297 (0.6%) | 354 (0.9%) | 207 (0.7%) | 237 (0.6%) | 1,095 (0.7%) |
| Asian | 291 (0.6%) | 279 (0.7%) | 338 (1.1%) | 375 (1.0%) | 1,283 (0.8%) |
| Chinese | 134 (0.3%) | 103 (0.3%) | 66 (0.2%) | 35 (0.1%) | 338 (0.2%) |
| Other | 209 (0.5%) | 306 (0.8%) | 126 (0.4%) | 205 (0.6%) | 846 (0.6%) |
| Missing[2] | 95 (0.2%) | 131 (0.3%) | 103 (0.3%) | 191 (0.5%) | 520 (0.3%) |
| Highest qualification | | | | | |
| None | 3,657 (7.9%) | 2,081 (5.2%) | 2,191 (7.4%) | 2,549 (6.9%) | 10,478 (6.9%) |
| O levels/GCSEs/CSEs | 12,694 (27.4%) | 9,163 (22.9%) | 6,024 (20.5%) | 7,904 (21.3%) | 35,785 (23.4%) |
| A levels/NVQ/HND/HNC[3] | 10,962 (23.6%) | 9,062 (22.7%) | 7,001 (23.8%) | 8,853 (23.9%) | 35,878 (23.5%) |
| Degree | 18,615 (40.2%) | 19,247 (48.2%) | 13,955 (47.4%) | 17,470 (47.1%) | 69,287 (45.3%) |
| Missing[2] | 433 (0.9%) | 397 (1.0%) | 274 (0.9%) | 331 (0.9%) | 1,435 (0.9%) |
| Household income[4] | | | | | |
| Very low | 6,004 (13.0%) | 5,748 (14.4%) | 2,974 (10.1%) | 4,149 (11.2%) | 18,875 (12.3%) |
| Low | 10,123 (21.8%) | 8,540 (21.4%) | 6,073 (20.6%) | 7,392 (19.9%) | 32,128 (21.0%) |
| Medium | 11,591 (25.0%) | 10,027 (25.1%) | 8,133 (27.6%) | 10,151 (27.4%) | 39,902 (26.1%) |
| High | 9,785 (21.1%) | 8,530 (21.4%) | 7,757 (26.3%) | 9,781 (26.4%) | 35,853 (23.5%) |
| Very high | 2,738 (5.9%) | 2,695 (6.7%) | 2,396 (8.1%) | 3,181 (8.6%) | 11,010 (7.2%) |
| Missing[2] | 6,120 (13.2%) | 4,410 (11.0%) | 2,112 (7.2%) | 2,453 (6.6%) | 15,095 (9.9%) |
| Neighbourhood deprivation | | | | | |
| Q1 | 11,146 (24.0%) | 7,860 (19.7%) | 7,563 (25.7%) | 7,999 (21.6%) | 34,568 (22.6%) |
| Q2 | 10,572 (22.8%) | 7,759 (19.4%) | 6,778 (23.0%) | 7,732 (20.8%) | 32,841 (21.5%) |
| Q3 | 9,867 (21.3%) | 7,976 (20.0%) | 6,108 (20.7%) | 7,472 (20.1%) | 31,423 (20.6%) |
| Q4 | 8,747 (18.9%) | 8,590 (21.5%) | 5,335 (18.1%) | 7,342 (19.8%) | 30,014 (19.6%) |
| Q5 | 5,975 (12.9%) | 7,714 (19.3%) | 3,624 (12.3%) | 6,510 (17.5%) | 23,823 (15.6%) |
| Missing[2] | 54 (0.1%) | 51 (0.1%) | 37 (0.1%) | 52 (0.1%) | 194 (0.1%) |

*Note:* Numbers shown are counts and percentages unless indicated otherwise. SD = standard deviation. GCSEs = general certificate of secondary education; CSE = certificate of secondary education; NVQ = national vocational qualification; HND = higher national diploma; HNC = higher national certificate. [1] Sample sizes for health and ageing markers: $n$ = 69,042 (MileAge delta), 4,252 (mortality profile), 152,689 (frailty index), 144,288 (T/S ratio) and 152,366 (grip strength). [2] Missing data may also include “do not know” or “prefer not to answer”. [3] Also includes 'other professional qualifications'. [4] Annual household income groups: very low (<£18,000), low (£18,000–£30,999), middle (£31,000–£51,999), high (£52,000–£100,000) and very high (>£100,000).

#### 7. Analytical sample sizes stratified by sex

**Table S7.** Non-relational trauma analytical sample sizes

| Non-relational trauma | MileAge delta | Mortality profile | Frailty index | Telomere length | Grip strength |
|---|---|---|---|---|---|
| Females | | | | | |
| Sum score[1] | 0.63 (0.85) | 0.61 (0.85) | 0.65 (0.86) | 0.64 (0.86) | 0.65 (0.86) |
| Sum score (sens.)[1] | 0.43 (0.68) | 0.43 (0.70) | 0.44 (0.69) | 0.44 (0.69) | 0.44 (0.69) |
| Specific items | | | | | |
| Combat | 38,916 (98.6%) | 2,440 (98.3%) | 87,461 (98.5%) | 82,410 (98.5%) | 87,267 (98.4%) |
| Yes | 560 (1.4%) | 42 (1.7%) | 1,376 (1.5%) | 1,296 (1.5%) | 1,375 (1.6%) |
| Accident | 36,809 (93.5%) | 2,306 (93.2%) | 82,645 (93.3%) | 77,875 (93.3%) | 82,460 (93.3%) |
| Yes | 2,559 (6.5%) | 167 (6.8%) | 5,961 (6.7%) | 5,603 (6.7%) | 5,950 (6.7%) |
| Illness | 33,541 (86.8%) | 2,099 (86.4%) | 75,118 (86.4%) | 70,883 (86.6%) | 74,954 (86.4%) |
| Yes | 5,081 (13.2%) | 330 (13.6%) | 11,777 (13.6%) | 10,984 (13.4%) | 11,751 (13.6%) |
| Crime | 33,825 (86.1%) | 2,164 (87.5%) | 75,860 (85.8%) | 71,499 (85.9%) | 75,706 (85.9%) |
| Yes | 5,464 (13.9%) | 309 (12.5%) | 12,507 (14.2%) | 11,763 (14.1%) | 12,465 (14.1%) |
| Sexual assault | 30,852 (79.6%) | 2,010 (82.4%) | 69,273 (79.4%) | 65,291 (79.4%) | 69,120 (79.4%) |
| Yes | 7,890 (20.4%) | 430 (17.6%) | 17,963 (20.6%) | 16,909 (20.6%) | 17,922 (20.6%) |
| Death witness | 35,921 (91.4%) | 2,258 (91.2%) | 80,738 (91.3%) | 76,090 (91.3%) | 80,565 (91.3%) |
| Yes | 3,371 (8.6%) | 218 (8.8%) | 7,680 (8.7%) | 7,219 (8.7%) | 7,658 (8.7%) |
| Males | | | | | |
| Sum score[1] | 0.83 (0.97) | 0.84 (0.96) | 0.85 (0.99) | 0.85 (0.98) | 0.85 (0.98) |
| Sum score (sens.)[1] | 0.76 (0.91) | 0.77 (0.90) | 0.78 (0.92) | 0.78 (0.92) | 0.78 (0.92) |
| Specific items | | | | | |
| Combat | 29,606 (94.2%) | 1,775 (94.7%) | 63,831 (93.9%) | 60,523 (93.9%) | 63,699 (93.9%) |
| Yes | 1,811 (5.8%) | 99 (5.3%) | 4,132 (6.1%) | 3,935 (6.1%) | 4,128 (6.1%) |
| Accident | 27,279 (87.0%) | 1,607 (86.1%) | 58,885 (86.8%) | 55,843 (86.8%) | 58,772 (86.8%) |
| Yes | 4,076 (13.0%) | 260 (13.9%) | 8,942 (13.2%) | 8,478 (13.2%) | 8,916 (13.2%) |
| Illness | 26,028 (84.9%) | 1,560 (85.4%) | 55,785 (84.3%) | 52,903 (84.3%) | 55,681 (84.3%) |
| Yes | 4,621 (15.1%) | 267 (14.6%) | 10,393 (15.7%) | 9,851 (15.7%) | 10,365 (15.7%) |
| Crime | 23,710 (75.8%) | 1,376 (73.5%) | 51,025 (75.4%) | 48,370 (75.4%) | 50,916 (75.4%) |
| Yes | 7,569 (24.2%) | 495 (26.5%) | 16,640 (24.6%) | 15,812 (24.6%) | 16,613 (24.6%) |
| Sexual assault | 28,973 (92.5%) | 1,735 (92.9%) | 62,577 (92.4%) | 59,359 (92.4%) | 62,449 (92.4%) |
| Yes | 2,343 (7.5%) | 132 (7.1%) | 5,131 (7.6%) | 4,863 (7.6%) | 5,121 (7.6%) |
| Death witness | 25,480 (81.6%) | 1,530 (82.2%) | 54,896 (81.3%) | 52,070 (81.3%) | 54,798 (81.3%) |
| Yes | 5,742 (18.4%) | 331 (17.8%) | 12,655 (18.7%) | 11,999 (18.7%) | 12,618 (18.7%) |

*Note:* Numbers shown are counts and percentages unless indicated otherwise. [1] mean and standard deviation.

#### 8. Sex-stratified non-relational trauma (sum scores)

**Table S8.** Sex-stratified associations between non-relational trauma and ageing markers

| | Model 1 (adj. age) | | | | Model 2 (full adjustment) | | | |
|---|---|---|---|---|---|---|---|---|
| Ageing marker | *β* | 95% CI | | *p* | *β* | 95% CI | | *p* |
| Females | | | | | | | | |
| MileAge delta | 0.024 | 0.012 | 0.036 | <0.001 | 0.024 | 0.011 | 0.036 | <0.001 |
| Metabolomic profile | 0.046 | 0.005 | 0.086 | 0.044 | 0.040 | 0.000 | 0.080 | 0.074 |
| Frailty index | 0.204 | 0.197 | 0.211 | <0.001 | 0.203 | 0.196 | 0.210 | <0.001 |
| Telomere length | 0.001 | -0.007 | 0.009 | 0.753 | 0.006 | -0.002 | 0.014 | 0.154 |
| Grip strength | 0.004 | -0.001 | 0.008 | 0.110 | 0.005 | 0.001 | 0.009 | 0.037 |
| Males | | | | | | | | |
| MileAge delta | 0.026 | 0.015 | 0.038 | <0.001 | 0.026 | 0.015 | 0.038 | <0.001 |
| Metabolomic profile | 0.057 | 0.017 | 0.098 | 0.010 | 0.043 | 0.003 | 0.084 | 0.053 |
| Frailty index | 0.176 | 0.169 | 0.182 | <0.001 | 0.167 | 0.160 | 0.173 | <0.001 |
| Telomere length | 0.006 | -0.002 | 0.013 | 0.203 | 0.005 | -0.003 | 0.013 | 0.227 |
| Grip strength | -0.003 | -0.009 | 0.003 | 0.326 | -0.009 | -0.015 | -0.003 | 0.005 |

*Note:* CI = confidence interval. Model 1–adjusted for chronological age; Model 2–adjusted for chronological age, ethnicity, highest educational/professional qualification, annual gross household income and neighbourhood deprivation. *P*-values shown are corrected for multiple testing using the Benjamini–Hochberg procedure (across ageing markers and models, separately for each sex). Sample sizes reported in Tables S7.

#### 9. Sex interactions non-relational trauma (sum scores)

**Table S9.** Sex interactions for associations between non-relational trauma and ageing markers

| | Model 1 (adj. age) | | | | Model 2 (full adjustment) | | | |
|---|---|---|---|---|---|---|---|---|
| | *β* | 95% CI | | *p* | *β* | 95% CI | | *p* |
| MileAge delta | 0.009 | -0.008 | 0.026 | 0.569 | 0.008 | -0.009 | 0.025 | 0.569 |
| Metabolomic profile | 0.007 | -0.050 | 0.065 | 0.892 | 0.001 | -0.056 | 0.058 | 0.968 |
| Frailty index | -0.028 | -0.037 | -0.018 | <0.001 | -0.035 | -0.044 | -0.025 | <0.001 |
| Telomere length | 0.002 | -0.009 | 0.013 | 0.853 | -0.002 | -0.013 | 0.009 | 0.853 |
| Grip strength | -0.007 | -0.014 | 0.000 | 0.146 | -0.007 | -0.014 | 0.000 | 0.146 |

*Note:* CI = confidence interval. Model 1–adjusted for chronological age; Model 2–adjusted for chronological age, ethnicity, highest educational/professional qualification, annual gross household income and neighbourhood deprivation. *P*-values shown are corrected for multiple testing using the Benjamini–Hochberg procedure (across ageing markers and models). Sample sizes reported in Table S7.

#### 10. Sex-stratified non-relational traumas (type-specific)

**Table S10.** Sex-stratified associations between non-relational trauma items and ageing markers

| Trauma exposure | Model 1 (adj. age) | | | | Model 2 (full adjustment) | | | |
|---|---|---|---|---|---|---|---|---|
| Females | | | | | | | | |
| | *β* | 95% CI | | *p* | *β* | 95% CI | | *p* |
| MileAge delta | | | | | | | | |
| Combat | 0.001 | -0.083 | 0.084 | 0.998 | -0.001 | -0.085 | 0.083 | 0.998 |
| Accident | 0.002 | -0.038 | 0.042 | 0.998 | 0.000 | -0.040 | 0.040 | 0.998 |
| Illness | 0.098 | 0.068 | 0.127 | <0.001 | 0.096 | 0.066 | 0.126 | <0.001 |
| Crime | 0.010 | -0.018 | 0.039 | 0.798 | 0.008 | -0.021 | 0.037 | 0.833 |
| Sexual assault | 0.036 | 0.011 | 0.060 | 0.015 | 0.034 | 0.008 | 0.059 | 0.022 |
| Death witness | 0.003 | -0.032 | 0.039 | 0.998 | 0.002 | -0.033 | 0.038 | 0.998 |
| Metabolomic profile | | | | | | | | |
| Combat | -0.071 | -0.331 | 0.190 | 0.833 | -0.023 | -0.282 | 0.236 | 0.998 |
| Accident | -0.041 | -0.176 | 0.094 | 0.833 | -0.031 | -0.165 | 0.102 | 0.841 |
| Illness | 0.194 | 0.095 | 0.293 | <0.001 | 0.183 | 0.085 | 0.281 | <0.001 |
| Crime | 0.016 | -0.086 | 0.118 | 0.950 | 0.003 | -0.098 | 0.104 | 0.998 |
| Sexual assault | 0.053 | -0.036 | 0.142 | 0.458 | 0.026 | -0.063 | 0.115 | 0.833 |
| Death witness | 0.000 | -0.118 | 0.119 | 0.998 | -0.004 | -0.121 | 0.114 | 0.998 |
| Frailty index | | | | | | | | |
| Combat | 0.138 | 0.090 | 0.186 | <0.001 | 0.132 | 0.085 | 0.180 | <0.001 |
| Accident | 0.259 | 0.236 | 0.283 | <0.001 | 0.261 | 0.238 | 0.285 | <0.001 |
| Illness | 0.397 | 0.379 | 0.414 | <0.001 | 0.390 | 0.374 | 0.407 | <0.001 |
| Crime | 0.186 | 0.169 | 0.203 | <0.001 | 0.172 | 0.155 | 0.189 | <0.001 |
| Sexual assault | 0.315 | 0.300 | 0.330 | <0.001 | 0.303 | 0.289 | 0.318 | <0.001 |
| Death witness | 0.211 | 0.190 | 0.232 | <0.001 | 0.218 | 0.197 | 0.238 | <0.001 |
| Telomere length | | | | | | | | |
| Combat | -0.047 | -0.100 | 0.007 | 0.197 | -0.025 | -0.079 | 0.028 | 0.640 |
| Accident | 0.009 | -0.017 | 0.035 | 0.821 | 0.019 | -0.008 | 0.045 | 0.331 |
| Illness | 0.001 | -0.019 | 0.020 | 0.998 | 0.002 | -0.018 | 0.021 | 0.998 |
| Crime | 0.005 | -0.014 | 0.024 | 0.833 | 0.013 | -0.006 | 0.033 | 0.331 |
| Sexual assault | 0.008 | -0.009 | 0.024 | 0.640 | 0.013 | -0.004 | 0.029 | 0.278 |
| Death witness | -0.016 | -0.040 | 0.007 | 0.331 | -0.009 | -0.033 | 0.014 | 0.741 |
| Grip strength | | | | | | | | |
| Combat | -0.006 | -0.035 | 0.023 | 0.845 | -0.008 | -0.037 | 0.021 | 0.833 |
| Accident | 0.012 | -0.003 | 0.026 | 0.246 | 0.017 | 0.002 | 0.031 | 0.053 |
| Illness | 0.025 | 0.014 | 0.035 | <0.001 | 0.025 | 0.015 | 0.036 | <0.001 |
| Crime | 0.014 | 0.004 | 0.024 | 0.021 | 0.014 | 0.004 | 0.025 | 0.018 |
| Sexual assault | 0.002 | -0.007 | 0.011 | 0.841 | 0.003 | -0.006 | 0.012 | 0.833 |
| Death witness | -0.029 | -0.042 | -0.016 | <0.001 | -0.025 | -0.037 | -0.012 | <0.001 |
| Males | | | | | | | | |
| MileAge delta | | | | | | | | |
| Combat | 0.024 | -0.023 | 0.070 | 0.500 | 0.022 | -0.025 | 0.069 | 0.500 |
| Accident | 0.015 | -0.017 | 0.048 | 0.500 | 0.014 | -0.018 | 0.047 | 0.531 |
| Illness | 0.152 | 0.121 | 0.183 | <0.001 | 0.150 | 0.119 | 0.181 | <0.001 |
| Crime | -0.007 | -0.033 | 0.018 | 0.680 | -0.007 | -0.033 | 0.019 | 0.680 |
| Sexual assault | 0.005 | -0.036 | 0.046 | 0.834 | 0.005 | -0.036 | 0.047 | 0.834 |
| Death witness | 0.028 | 0.000 | 0.056 | 0.116 | 0.027 | -0.002 | 0.055 | 0.132 |
| Metabolomic profile | | | | | | | | |
| Combat | 0.134 | -0.038 | 0.305 | 0.245 | 0.069 | -0.101 | 0.239 | 0.546 |
| Accident | -0.052 | -0.164 | 0.059 | 0.500 | -0.054 | -0.164 | 0.057 | 0.500 |
| Illness | 0.288 | 0.178 | 0.399 | <0.001 | 0.275 | 0.165 | 0.384 | <0.001 |
| Crime | 0.044 | -0.044 | 0.133 | 0.500 | 0.027 | -0.061 | 0.115 | 0.680 |

| | | | | | | | | |
|---|---|---|---|---|---|---|---|---|
| Sexual assault | 0.028 | -0.121 | 0.178 | 0.760 | -0.016 | -0.165 | 0.133 | 0.844 |
| Death witness | 0.054 | -0.047 | 0.154 | 0.488 | 0.027 | -0.073 | 0.127 | 0.680 |
| Frailty index | | | | | | | | |
| Combat | 0.162 | 0.134 | 0.190 | <0.001 | 0.125 | 0.098 | 0.153 | <0.001 |
| Accident | 0.259 | 0.239 | 0.278 | <0.001 | 0.245 | 0.226 | 0.264 | <0.001 |
| Illness | 0.479 | 0.461 | 0.497 | <0.001 | 0.460 | 0.442 | 0.478 | <0.001 |
| Crime | 0.184 | 0.168 | 0.199 | <0.001 | 0.171 | 0.155 | 0.186 | <0.001 |
| Sexual assault | 0.279 | 0.254 | 0.304 | <0.001 | 0.259 | 0.235 | 0.284 | <0.001 |
| Death witness | 0.161 | 0.144 | 0.178 | <0.001 | 0.153 | 0.136 | 0.169 | <0.001 |
| Telomere length | | | | | | | | |
| Combat | 0.006 | -0.025 | 0.037 | 0.760 | 0.003 | -0.029 | 0.034 | 0.870 |
| Accident | 0.015 | -0.008 | 0.037 | 0.338 | 0.015 | -0.007 | 0.037 | 0.319 |
| Illness | 0.017 | -0.004 | 0.038 | 0.213 | 0.016 | -0.005 | 0.037 | 0.247 |
| Crime | 0.007 | -0.010 | 0.025 | 0.546 | 0.009 | -0.009 | 0.026 | 0.500 |
| Sexual assault | 0.008 | -0.020 | 0.036 | 0.680 | 0.008 | -0.021 | 0.036 | 0.680 |
| Death witness | -0.005 | -0.024 | 0.014 | 0.685 | -0.008 | -0.027 | 0.011 | 0.546 |
| Grip strength | | | | | | | | |
| Combat | -0.055 | -0.079 | -0.031 | <0.001 | -0.070 | -0.094 | -0.046 | <0.001 |
| Accident | -0.028 | -0.045 | -0.011 | 0.003 | -0.038 | -0.055 | -0.021 | <0.001 |
| Illness | 0.042 | 0.026 | 0.058 | <0.001 | 0.036 | 0.020 | 0.052 | <0.001 |
| Crime | 0.049 | 0.035 | 0.063 | <0.001 | 0.034 | 0.021 | 0.048 | <0.001 |
| Sexual assault | 0.031 | 0.009 | 0.052 | 0.013 | 0.016 | -0.005 | 0.038 | 0.258 |
| Death witness | -0.086 | -0.100 | -0.071 | <0.001 | -0.085 | -0.099 | -0.070 | <0.001 |

*Note:* CI = confidence interval. Model 1–adjusted for chronological age; Model 2–adjusted for chronological age, ethnicity, highest educational/professional qualification, annual gross household income and neighbourhood deprivation. *P*-values shown are corrected for multiple testing using the Benjamini–Hochberg procedure (across exposures, ageing markers and models, separately for each sex). Sample sizes reported in Table S7.

#### 11. Sex interactions non-relational traumas (type-specific)

**Table S11.** Sex interactions for associations between non-relational trauma items and ageing markers

| Trauma exposure | Model 1 (adj. age) | | | | Model 2 (full adjustment) | | | |
|---|---|---|---|---|---|---|---|---|
| | *β* | 95% CI | | *p* | *β* | 95% CI | | *p* |
| **MileAge delta** | | | | | | | | |
| Combat | 0.003 | -0.092 | 0.099 | >0.999 | 0.000 | -0.096 | 0.096 | >0.999 |
| Accident | 0.034 | -0.017 | 0.086 | 0.384 | 0.032 | -0.019 | 0.084 | 0.425 |
| Illness | 0.000 | -0.043 | 0.043 | >0.999 | 0.000 | -0.043 | 0.043 | >0.999 |
| Crime | 0.029 | -0.009 | 0.068 | 0.313 | 0.028 | -0.010 | 0.066 | 0.330 |
| Sexual assault | -0.017 | -0.066 | 0.031 | 0.784 | -0.017 | -0.066 | 0.032 | 0.784 |
| Death witness | 0.036 | -0.009 | 0.081 | 0.305 | 0.035 | -0.010 | 0.080 | 0.311 |
| **Metabolomic profile** | | | | | | | | |
| Combat | 0.218 | -0.094 | 0.529 | 0.353 | 0.131 | -0.177 | 0.439 | 0.689 |
| Accident | -0.030 | -0.204 | 0.145 | 0.944 | -0.044 | -0.216 | 0.128 | 0.882 |
| Illness | 0.116 | -0.031 | 0.264 | 0.305 | 0.117 | -0.029 | 0.262 | 0.305 |
| Crime | 0.002 | -0.132 | 0.136 | >0.999 | -0.004 | -0.136 | 0.128 | >0.999 |
| Sexual assault | -0.027 | -0.203 | 0.148 | 0.948 | -0.037 | -0.210 | 0.136 | 0.900 |
| Death witness | 0.050 | -0.106 | 0.205 | 0.817 | 0.033 | -0.121 | 0.186 | 0.900 |
| **Frailty index** | | | | | | | | |
| Combat | 0.023 | -0.032 | 0.078 | 0.689 | -0.007 | -0.061 | 0.047 | 0.958 |
| Accident | 0.000 | -0.031 | 0.031 | >0.999 | -0.017 | -0.047 | 0.013 | 0.487 |
| Illness | 0.076 | 0.051 | 0.101 | <0.001 | 0.068 | 0.044 | 0.092 | <0.001 |
| Crime | -0.003 | -0.026 | 0.020 | 0.956 | -0.005 | -0.027 | 0.018 | 0.900 |
| Sexual assault | -0.035 | -0.064 | -0.006 | 0.079 | -0.040 | -0.068 | -0.011 | 0.034 |
| Death witness | -0.050 | -0.077 | -0.023 | 0.002 | -0.065 | -0.091 | -0.038 | <0.001 |
| **Telomere length** | | | | | | | | |
| Combat | 0.058 | -0.004 | 0.120 | 0.188 | 0.037 | -0.025 | 0.099 | 0.451 |
| Accident | 0.000 | -0.034 | 0.035 | >0.999 | -0.007 | -0.042 | 0.027 | 0.900 |
| Illness | 0.032 | 0.003 | 0.061 | 0.099 | 0.030 | 0.002 | 0.059 | 0.130 |
| Crime | -0.012 | -0.038 | 0.013 | 0.603 | -0.019 | -0.045 | 0.007 | 0.324 |
| Sexual assault | -0.003 | -0.036 | 0.029 | 0.990 | -0.008 | -0.041 | 0.024 | 0.882 |
| Death witness | 0.009 | -0.022 | 0.039 | 0.863 | 0.000 | -0.031 | 0.030 | >0.999 |
| **Grip strength** | | | | | | | | |
| Combat | -0.048 | -0.088 | -0.008 | 0.079 | -0.045 | -0.085 | -0.005 | 0.099 |
| Accident | -0.040 | -0.063 | -0.018 | 0.002 | -0.043 | -0.065 | -0.021 | 0.001 |
| Illness | 0.019 | 0.000 | 0.037 | 0.141 | 0.019 | 0.000 | 0.037 | 0.141 |
| Crime | 0.032 | 0.015 | 0.049 | 0.001 | 0.033 | 0.017 | 0.050 | <0.001 |
| Sexual assault | 0.028 | 0.007 | 0.049 | 0.051 | 0.026 | 0.004 | 0.047 | 0.079 |
| Death witness | -0.057 | -0.076 | -0.037 | <0.001 | -0.058 | -0.078 | -0.039 | <0.001 |

*Note:* CI = confidence interval. Model 1–adjusted for chronological age; Model 2–adjusted for chronological age, ethnicity, highest educational/professional qualification, annual gross household income and neighbourhood deprivation. *P*-values shown are corrected for multiple testing using the Benjamini–Hochberg procedure (across exposures, ageing markers and models). Sample sizes reported in Table S7.

12. Non-relational traumas (sum scores, excluding sexual assault)

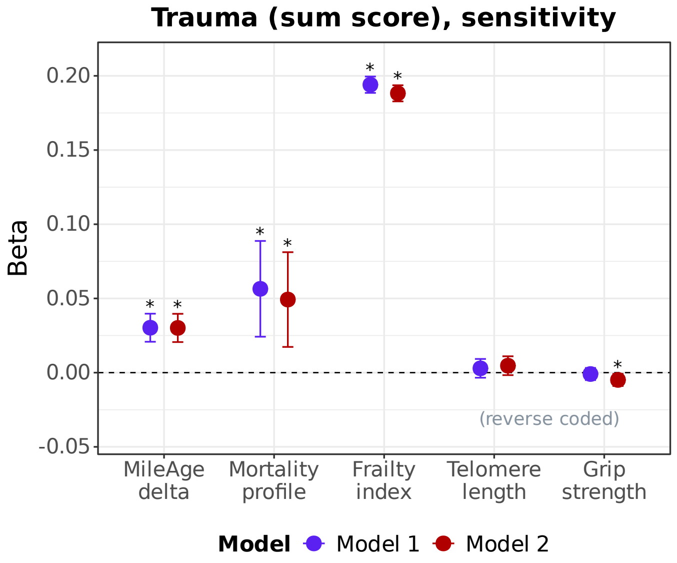

**Figure S2. Trauma (sum scores, excluding sexual assault) and ageing markers.** Associations between lifetime exposure to non-relational trauma and ageing markers (MileAge delta, frailty index, telomere length [reverse coded] and grip strength [reverse coded]). Estimates shown are ordinary least squares regression beta coefficients and 95% confidence intervals. Model 1–adjusted for chronological age and sex; Model 2–adjusted for chronological age, sex, ethnicity, education, income and neighbourhood deprivation. Asterisks indicate statistically significant associations, after correcting p-values for multiple testing using the Benjamini–Hochberg procedure (across ageing markers and models). Sample sizes reported in Table S2.

**Table S12.** Associations between non-relational trauma and ageing markers (sum score, excluding sexual assault)

| | Model 1 (adj. age and sex) | | | | Model 2 (full adjustment) | | | |
|---|---|---|---|---|---|---|---|---|
| Ageing marker | *β* | 95% CI | | *p* | *β* | 95% CI | | *p* |
| Non-relational trauma | | | | | | | | |
| MileAge delta | 0.030 | 0.021 | 0.040 | <0.001 | 0.030 | 0.021 | 0.040 | <0.001 |
| Metabolomic profile | 0.056 | 0.024 | 0.089 | 0.001 | 0.049 | 0.017 | 0.081 | 0.004 |
| Frailty index | 0.194 | 0.189 | 0.200 | <0.001 | 0.188 | 0.183 | 0.194 | <0.001 |
| Telomere length | 0.003 | -0.003 | 0.009 | 0.408 | 0.005 | -0.002 | 0.011 | 0.186 |
| Grip strength | -0.001 | -0.005 | 0.003 | 0.660 | -0.005 | -0.009 | -0.001 | 0.027 |

*Note:* CI = confidence interval. Model 1–adjusted for chronological age and sex; Model 2–adjusted for chronological age, sex, ethnicity, highest educational/professional qualification, annual gross household income and Townsend deprivation index. *P*-values shown are corrected for multiple testing using the Benjamini–Hochberg procedure (across ageing markers and models). Sample sizes reported in Table S2.

13. Sex-stratified non-relational trauma (sum scores, excluding sexual assault)

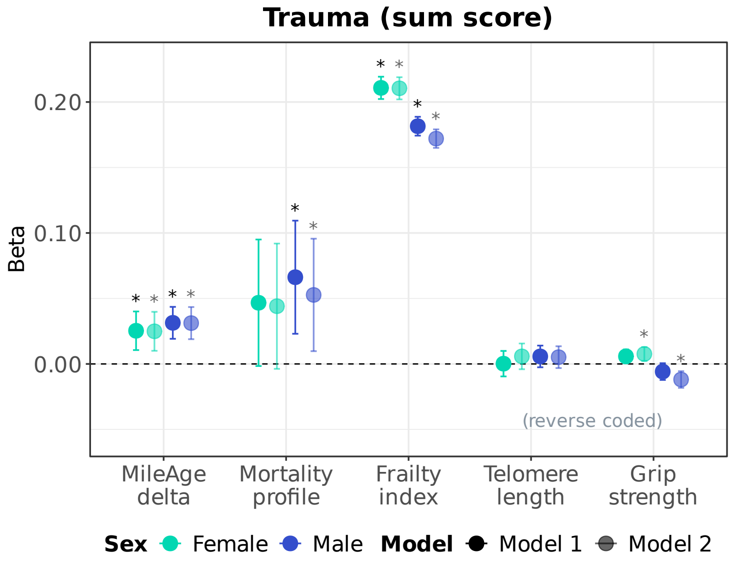

**Figure S3. Sex-stratified associations between trauma (sum scores, excluding sexual assault) and ageing markers.** Sex-stratified associations between lifetime exposure to non-relational trauma and ageing markers (MileAge delta, frailty index, telomere length [reverse coded] and grip strength [reverse coded]). Estimates shown are ordinary least squares regression beta coefficients and 95% confidence intervals. Model 1–adjusted for chronological age; Model 2–adjusted for chronological age, ethnicity, highest educational/professional qualification, annual gross household income and neighbourhood deprivation. Asterisks indicate statistically significant associations, after correcting *p*-values for multiple testing using the Benjamini–Hochberg procedure (across ageing markers and models, separately for each sex). Sample sizes reported in Table S7.

**Table S13.** Sex-stratified associations between non-relational trauma types (excluding sexual assault) and ageing markers

| | Model 1 (adj. age) | | | | Model 2 (full adjustment) | | | |
|---|---|---|---|---|---|---|---|---|
| Ageing marker | *β* | 95% CI | | *p* | *β* | 95% CI | | *p* |
| Females | | | | | | | | |
| MileAge delta | 0.025 | 0.011 | 0.040 | 0.003 | 0.025 | 0.010 | 0.040 | 0.003 |
| Metabolomic profile | 0.047 | -0.002 | 0.095 | 0.083 | 0.044 | -0.004 | 0.092 | 0.089 |
| Frailty index | 0.211 | 0.202 | 0.219 | <0.001 | 0.210 | 0.202 | 0.219 | <0.001 |
| Telomere length | 0.000 | -0.010 | 0.010 | 0.968 | 0.006 | -0.004 | 0.016 | 0.277 |
| Grip strength | 0.006 | 0.001 | 0.011 | 0.052 | 0.008 | 0.002 | 0.013 | 0.009 |
| Males | | | | | | | | |
| MileAge delta | 0.031 | 0.019 | 0.044 | <0.001 | 0.031 | 0.019 | 0.043 | <0.001 |
| Metabolomic profile | 0.066 | 0.023 | 0.109 | 0.004 | 0.053 | 0.010 | 0.096 | 0.023 |
| Frailty index | 0.182 | 0.174 | 0.189 | <0.001 | 0.172 | 0.165 | 0.179 | <0.001 |
| Telomere length | 0.006 | -0.003 | 0.014 | 0.195 | 0.005 | -0.003 | 0.014 | 0.223 |
| Grip strength | -0.006 | -0.012 | 0.001 | 0.092 | -0.012 | -0.018 | -0.006 | <0.001 |

*Note:* CI = confidence interval. Model 1–adjusted for chronological age; Model 2–adjusted for chronological age, ethnicity, highest educational/professional qualification, annual gross household income and neighbourhood deprivation. *P*-values shown are corrected for multiple testing using the Benjamini–Hochberg procedure (across ageing markers and models, separately for each sex). Sample sizes reported in Tables S7.

#### 14. Sex interactions non-relational trauma (sum scores, excluding sexual assault)

**Table S14.** Sex interactions for associations between non-relational trauma (excluding sexual assault) and ageing markers

| | Model 1 (adj. age) | | | | Model 2 (full adjustment) | | | |
|---|---|---|---|---|---|---|---|---|
| Ageing marker | *β* | 95% CI | | *p* | *β* | 95% CI | | *p* |
| MileAge delta | 0.011 | -0.009 | 0.030 | 0.552 | 0.010 | -0.010 | 0.029 | 0.552 |
| Metabolomic profile | 0.015 | -0.049 | 0.080 | 0.802 | 0.007 | -0.057 | 0.071 | 0.856 |
| Frailty index | -0.029 | -0.040 | -0.018 | <0.001 | -0.038 | -0.049 | -0.027 | <0.001 |
| Telomere length | 0.004 | -0.009 | 0.017 | 0.732 | -0.001 | -0.014 | 0.012 | 0.856 |
| Grip strength | -0.012 | -0.020 | -0.003 | 0.013 | -0.012 | -0.020 | -0.004 | 0.013 |

*Note:* CI = confidence interval. Model 1–adjusted for chronological age; Model 2–adjusted for chronological age, ethnicity, highest educational/professional qualification, annual gross household income and neighbourhood deprivation. *P*-values shown are corrected for multiple testing using the Benjamini–Hochberg procedure (across ageing markers and models). Sample sizes reported in Table S7.
